# The public health co-benefits of strategies consistent with net-zero emissions: a systematic review of quantitative studies

**DOI:** 10.1101/2024.08.26.24312597

**Authors:** Léo Moutet, Paquito Bernard, Rosemary Green, James Milner, Andy Haines, Rémy Slama, Laura Temime, Kévin Jean

**Author notes:** Author to whom any correspondence should be addressed.

## Abstract

Moving toward net-zero emission societies is projected to provide health co-benefits, yet their magnitude is not well documented and may be context-specific. Synthesizing the evidence on these co-benefits could enhance the engagement of decision-makers and populations in climate mitigation actions. We performed a systematic review including 58 quantitative studies exploring 125 scenarios. Across air quality, physical activity and dietary changes pathways, substantial health co-benefits were found, with half of scenarios showing a mortality reduction by more than 1.5%, in addition to benefits directly related to climate stabilization. However, these co-benefits varied with explored emission sectors, decarbonization levers, modelling approaches and locations. Among studies including a cost-benefit analysis, 11 of 13 estimated that monetized benefits outweighed the costs of implementing climate policies. This review highlights the need for a standardised framework to assess and compare health impacts of climate mitigation actions across sectors, and confirms that achieving net-zero goals supports far-reaching public health policies.

## INTRODUCTION

In 2016, 196 governments signed the Paris agreement that aims to reduce anthropogenic greenhouse gas (GHG) emissions to net-zero by mid-century to limit global warming well below 2°C above preindustrial levels.^1^ Resulting nationwide commitments, identified as Nationally Determined Contributions (NDCs), fall short of addressing these objectives and a majority of currently implemented policies do not achieve pledged contributions.^2,3^ Beyond NDCs, various governmental or non-governmental organizations have been developing roadmaps that outline technical and political solutions for society to attain net-zero emissions (i.e. GHG emissions reduced to the lowest possible level with remaining emissions being offset by natural or artificial carbon sinks). These strategies activate different levers, such as technological innovation improving energy efficiency and allowing decarbonized energy production; or political, fiscal and behavioural instruments, reducing the use of energy and materials, often referred to as demand-side policies.

Many climate mitigation policies are likely to also benefit human health by directly and indirectly targeting modifiable environmental and behavioural risks, such as air pollution or diet.^2,4^ Several studies have assessed the health co-benefits arising from either single climate mitigation actions or regional or national multi-sectoral climate policies.^5,6^ Recently, the Lancet Pathfinder initiative produced an umbrella review exploring the health co-benefits of a wide range of specific GHG mitigation actions.^4^ As yet, no systematic review has explored the health impact of combinations of actions aimed at achieving net-zero emissions.

Such an appraisal could provide valuable insights for identifying specific health pathways, sectors of activity or levers of decarbonization that are likely to optimize the co-benefits of climate mitigation actions. Summarizing the existing evidence regarding the health co-benefits of pathways to net-zero GHG emissions is also key to increasing the commitment of people and their governments to climate actions in a context where implemented or pledged policies fall short of the goals of the Paris Agreement.^7,8^

Here, we systematically reviewed the current evidence regarding the health co-benefits of prospective net-zero GHG emission scenarios (thereafter “net-zero scenarios”). We compare the predicted health co-benefits across published health impact assessment (HIA) studies, accounting for various sectors of activity and co-benefit pathways. We also identify the main gaps in knowledge, needs for future research, and provide some recommendations for health impact assessments of prospective net-zero emission scenarios.

## METHODS

We conducted a systematic review, following the Preferred Reporting Items for Systematic Review and Meta-analysis (PRISMA) 2020 guidelines^9^. The PRISMA checklist is available as Table S1. The study protocol was registered in PROSPERO (ID: CRD42023429759).

### Search strategy

We searched three literature databases for studies published prior to January 2024: PubMed, Web of Science and Scopus. The search query included two mandatory terms, referring to health or mortality on the one side; and to net-zero emissions targets or limited climate change on the other. The detailed strategy is available in Table S2.

### Selection criteria and screening

Studies were screened by two independent reviewers (LM and KJ) using the Covidence management tool.^10^ A third researcher (LT) resolved any conflicts.

Screening was first carried out based on titles and abstracts (step 1), from which only original research pieces were included. At this stage, we only included studies explicitly referring to a GHG emission objective and assessing quantitative health outcomes or an economic valuation of health impacts. Qualitative studies, reviews, meta-analyses or opinion pieces were excluded although we screened meta-analyses and reviews for potential studies to include.

In the full-text assessment (step 2), we included studies which: 1) relied on a prospective scenario that included socio-economic and/or technical choices sufficient to attain net-zero GHG emissions or meet Paris agreement objectives (a climate warming limited to 1.5 °C or failing that to well under 2°C); 2) provided quantitative estimates of health impacts or economic assessments of such benefits; and 3) explored at least one health co-benefit pathway of mitigation actions. The studies were not required to assess all health pathways that would be affected by the emission sectors considered in the overall prospective scenario.

Co-benefits pathways were defined here as climate mitigation actions that improve human health by pathways, unmediated by climate. They included, but were not *a priori* limited to, air quality improvement, enhanced active transport and healthy dietary patterns. We considered the mitigation of extreme heat or extreme climatic events as a direct benefit of climate mitigation policies; and therefore excluded them from quantitative analyses.

### Data extraction

For all included articles, two authors (LM and PB) independently extracted information on the following characteristics: time and geographical scale, emission sector(s) considered (power generation, transportation, agriculture), explored co-benefits pathways (e.g. diet, physical activity, air pollution…) and assessed health outcome metrics (number of deaths prevented, life-years gained…). When available, the disaggregated impacts estimated across different sectors or pathways were extracted. We also retrieved characteristics regarding the modelling methods: demographic hypothesis, models of exposure, health impact assessment approach, and exposure-response function applied.

For each study (and each scenario assessed when the study assessed several), we categorized net-zero scenarios based on the major lever of mitigation assumed, using the following in-house categorisation: energy decarbonization, demand reduction, health in climate policies, financial instrument. Baseline scenarios were also categorized based on their assumptions regarding evolution of GHG emissions or utilization of a reference year (Figure S1).

### Confidence assessment

Since there is no validated tool to assess methodological bias in health impact assessment studies, we referred to guidelines reported by Hess et al for modelling and reporting health effects of climate change mitigation actions.^11^ Among 36 modelling and/or reporting criteria suggested by Hess et al, we retrieved those relevant to our study context and merged them into major topics, ending up with 13 final criteria (see table S3 for details).

### Health impacts scaling

In order to compare health impacts across studies, we retrieved and scaled estimates of the number of deaths prevented and/or life-years gained. When only life-years gained were estimated and if the region of investigation was available in the Global Burden of Disease 2021, they were converted into premature deaths prevented.^12^ The scaled outcome analysed was the preventable mortality fraction, estimated based on the ratio between the number of deaths prevented by a scenario relative to a baseline and the number of deaths projected for the associated location, time and age range. More details on the scaling calculations are provided in supplementary text 1. Analyses were conducted using R and are available at: https://github.com/LeoMoutet/revue_syst.

## RESULTS

### Descriptive findings

We identified 3,976 records from the three databases, of which 1,433 duplicates were removed (Figure 1). Of the 2,582 abstracts screened (step 1), 92 qualified for full-text screening. In the full-text assessment (step 2), 34 studies were excluded, mainly because they did not estimate quantitative health metrics (n=10) or because they were not explicitly based on net-zero scenarios (n=14). All corresponding authors from included studies were contacted in December 2023 to request potential relevant unidentified peer-reviewed studies, resulting in the inclusion of two additional studies. Eventually, 58 studies met our inclusion criteria.

**Figure 1.**
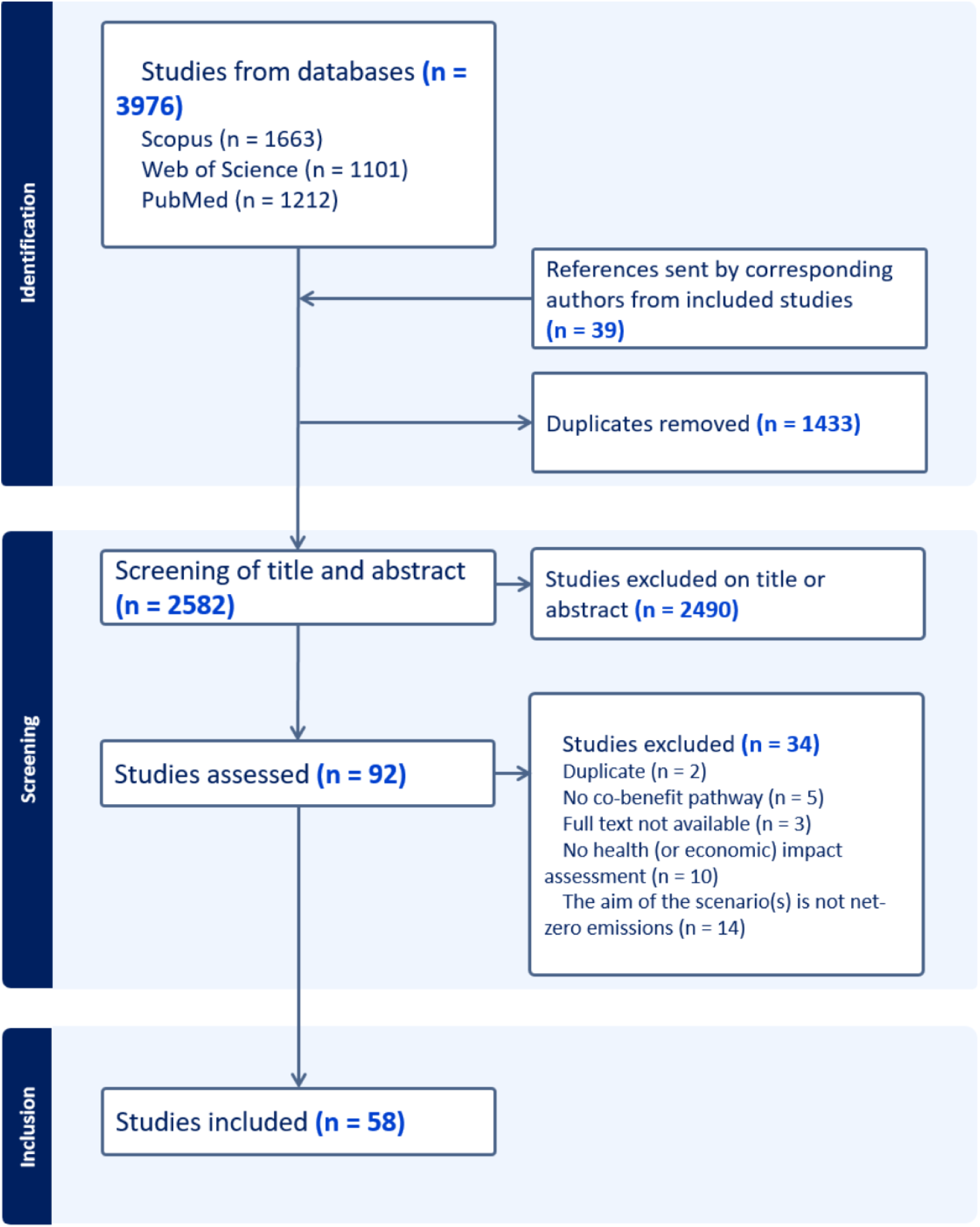
Flow-chart of study selection.

In addition to 12 worldwide studies,^13–24^ eight were conducted on a multinational scale (Figure 2) involving from two to 139 countries^5,6,25–30^, and 25 on single countries. These national assessments focused on north-east Asia,^31–48^ Europe,^49–52^ India^53,54^or the USA^55^ and 13 sub-national studies conducted in east-China,^56–61^ Europe,^62,63^ California (USA),^64–66^ Virginia (USA),^67^ and Santiago de Chile.^68^

**Figure 2.**
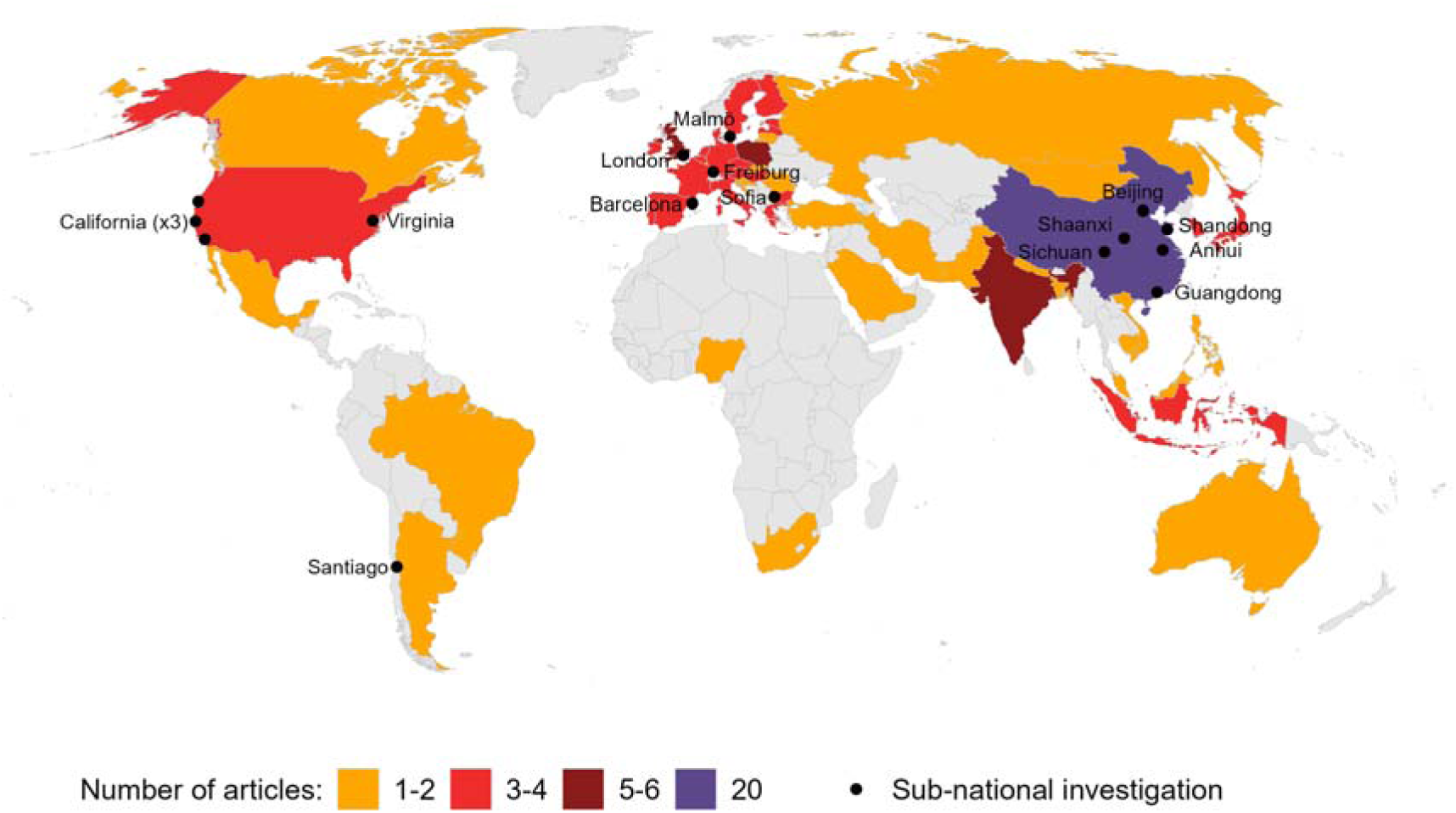
Geographical distribution of studies included. *Worldwide studies (n=12) are not represented on the map*.

The main characteristics of included studies are described in Figure 3. The majority (91%) of the included papers were published since 2018 (Figure 3A).

**Figure 3.**
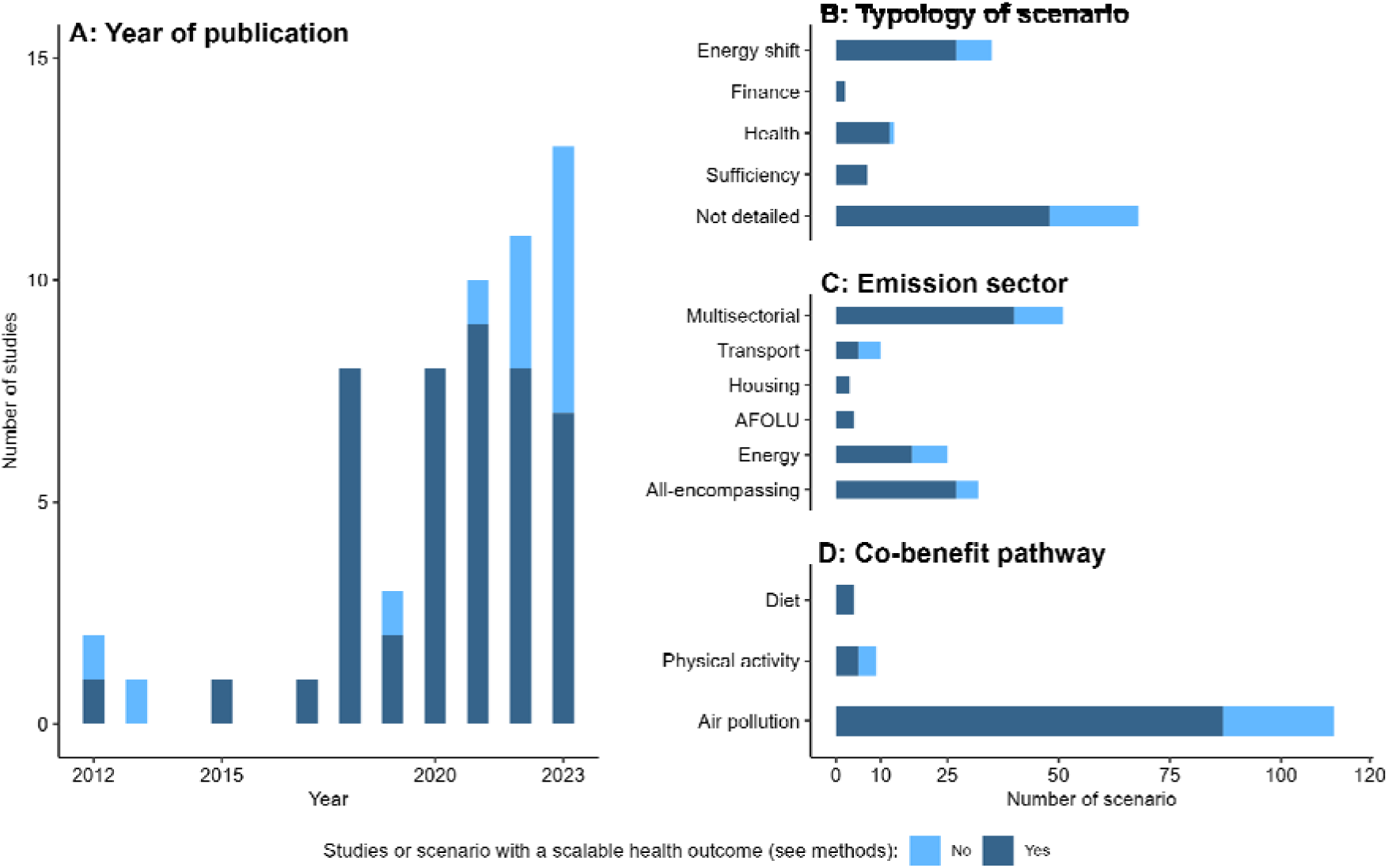
Descriptive analysis of included studies, by publication year (A), type of scenario (B), emission sector (C) and co-benefit pathway studied (D).

### Net-zero emission scenarios

14 studies assessed comprehensive scenarios from external prospective net-zero emission plans, i.e. developed by a governmental or non-governmental institution. Ten studies based their scenarios on official NDCs and 20 studies relied on the temperature target from the Paris agreement to estimate subsequent GHG emissions and air pollution projections. For 14 studies, the authors developed an in-house scenario (e.g. Net-zero CO2 emission target year for each G20 countries) to assess the impacts of various specific measures (more details in supplementary text 2).

Out of 125 scenarios, 58 provided specific details on the projected levers to achieve net-zero emissions (Figure 3B). The main policy lever identified was decarbonization of the energy sector through the scale-up of technologies such as carbon capture and storage, renewable energy, electrification or development of nuclear energy production. Some scenarios aimed specifically at the improvement of human health in a “health in all policies” approach, most commonly by improving air quality.^5,18,19,21,27,29,38,53,57,62,66,68^ A few scenarios relied on demand-side interventions (e.g. decreased energy or transport demand, n=7)^6,17,31,35,47,52,56^ or financial instruments (e.g. carbon taxes or prices of parking, n=4)^16,53,62,66^, projected to induce various behavioural shifts (Figure S1).

### Emission sectors and co-benefit pathways considered

Heterogenous combinations of emission sectors and co-benefits pathways were explored (Figure 4). The emission sectors most frequently studied were energy (n=40), transport (n=27), industry (n=21), housing (n=15) and Agriculture, Forestry and Other Land Use (AFOLU) (n=13) (Figure 3C). A majority (n=23) of the studies were multi-sectoral and 14 studies modelled global anthropogenic emissions, with some studies including natural emissions (such as vegetation fire, dust, sea sprays, biogenic volatile organic compounds…). These models do not incorporate any specific changes in natural emissions based on the scenarios. The vast majority of studies (n=56) assessed health impacts related to air quality, including fine particulate matter or PM_2.5_ (n=53), O_3_ (n=22), SO_2_ (n=4), NO_x_ (n=3), NO_2_ (n=4), and PM_10_ (n=3); five of these included household exposures to PM_2.5_ (n=5), radon and tobacco smoke (n=2), O_3_ (n=1), increased winter temperature attributable to home energy efficiency (n=1) and mould (n=1). Out of the studies including PM_2.5_, 17 considered specifically black carbon. Six scenarios investigated physical activity enhanced by active transport, while five scenarios examined dietary changes, with notably a reduction in red meat consumption (Figure 3D). Two studies combined air pollution, diet and physical activity,^5,6^ two studies focused exclusively on physical activity ^52,62^ and one on household air temperature and air quality (PM, radon, tobacco smoke and mould).^63^

**Figure 4.**
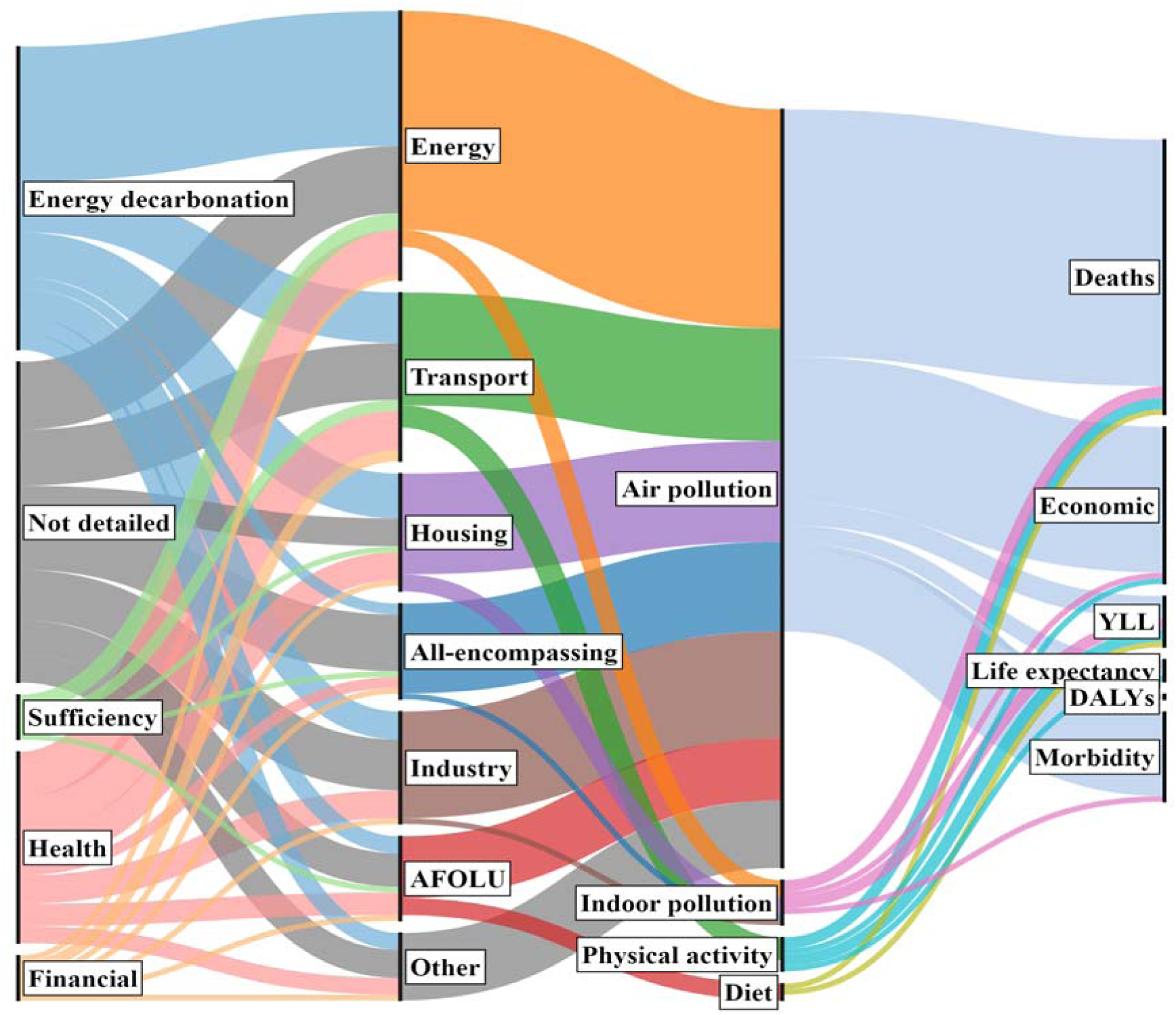
Linkage between typology of scenario, sector of emission, co-benefit pathway and health outcome across net-zero scenarios. *Each scenario can have links to several emissions sectors, exposition and outcome. AFOLU: Agriculture, Forestry and Other Land Use*.

### Methods used

Various health outcomes were quantified in the studies selected: 46 estimated the number of premature deaths prevented, four calculated changes in life expectancy, six assessed life-years gained and one calculated disability-adjusted life years. Additionally, seven studies specified morbidity outcomes and 28 studies conducted an economic assessment, mainly using the value of a statistical life year (n=24), with some studies adding a cost of illness (n=5) or a social cost of carbon (n=2) assessment. Others based their assessment on external costs from the European Commission (n=2), the unit value of health outcome (n=1) or the cost of conserved energy (n=1).

Several framework for modelling exposure were used across included studies to: 1) spatialize air pollution concentrations based on emissions reduction using a single model or a model mixture (atmospheric-chemistry, energy system, integrated assessment with air quality module); 2) attribute health outcomes to changes in active transport in the population; 3) attribute health outcomes to changes in dietary patterns in the population.

Methods to quantify health impacts were more limited in number, with 44 studies using comparative risk assessment methods (CRA), 13 studies relying on lifetable approaches, and one employing microsimulations.^53^

### Confidence assessment

According to our criteria adapted from Hess et al.^11^, general modelling methods were overall well conducted (Figure 5, criteria 1 to 6). The policies, scenarios and timeframes were well defined, whereas the most overlooked criterion was the evaluation of the equity impacts of policy adoption. Discussion of the adverse consequences of mitigation actions, sources of uncertainty and sensitivity analyses were limited. There were also very little data and code publicly available. Detailed results of the confidence assessment by study are available in Table S4.

**Figure 5.**
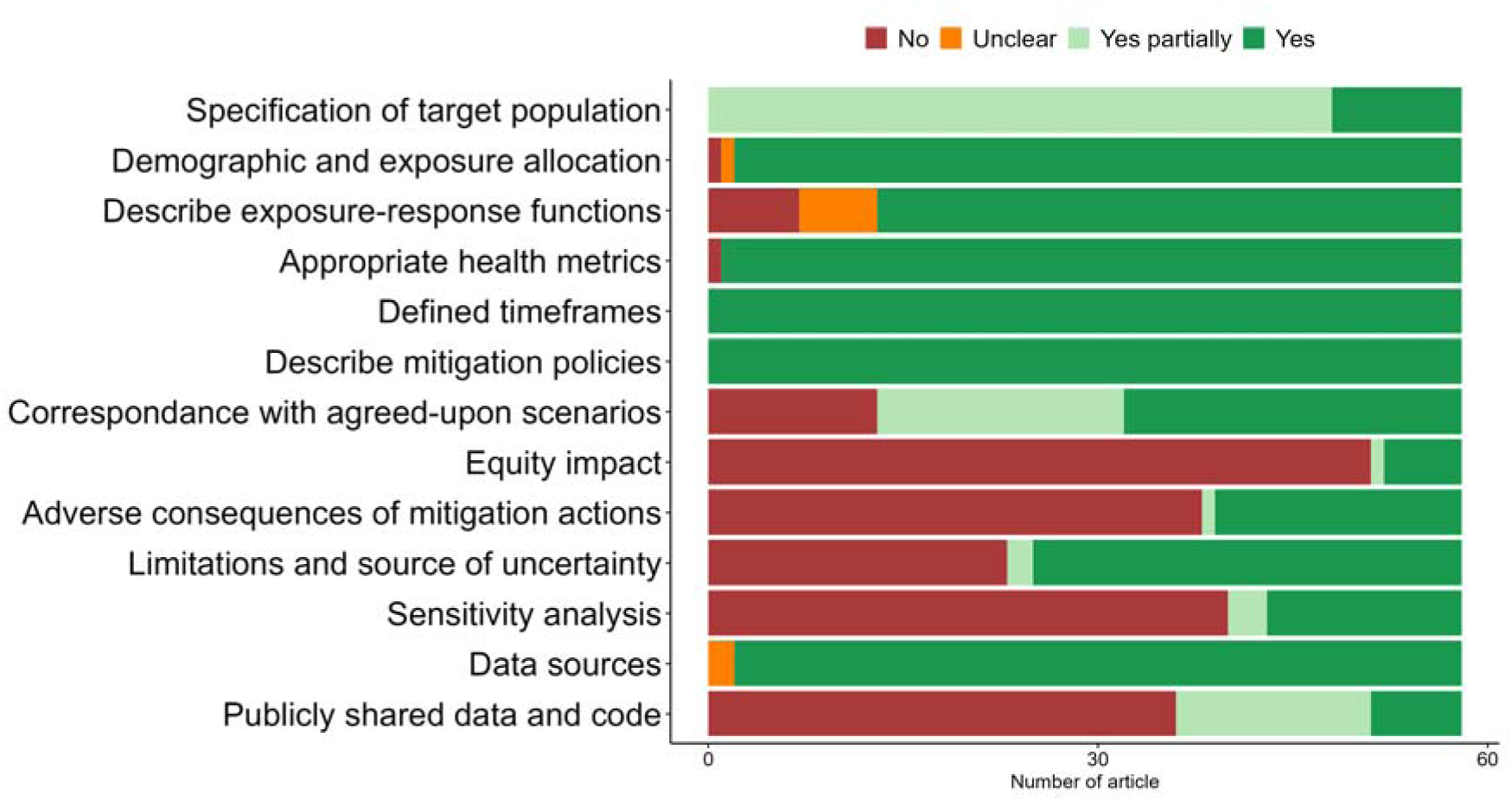
Confidence assessment of included studies per criterion adapted from Hess et al.^11^.

### Synthesis of the evidence

#### Quantitative health impact

We were able to retrieve and scale the preventable mortality fraction of 96 scenarios across 45 studies. Across these scenarios, two (from one study) reported detrimental health impacts (i.e. adverse effects on health) in the energy sector (–0.09% and –0.04% of mortality fraction).^51^ All other scenarios (i.e. 94 over 96) yielded considerable reductions in all-cause mortality, with an inter-quartile range between 0.55% and 3.59%, and up to 18.74% (highest estimated impact),^45^ with a median value of 1.48% (Figure 6A). The estimated health impacts seemed lower in studies using lifetables and higher when accounting for increasing GHG emissions in the baseline scenario (Figure 6 B/C), a finding which holds true even when considering air pollution pathway only (Figure S2). Although very few studies assessed the impacts of diet and physical activity pathways, the benefits arising from changing their patterns have the potential to yield significant health benefits (Figure 6D). Modelling emissions from multiple or unique sectors may have provided as much health benefit compared with using whole economy models (Figure 6E). We did not identify any single common factor among the scenarios that yielded the greatest health benefits. When comparing the economic benefits arising from health impacts and the implementation costs of the policies (n=13), most studies (n=11) found net benefits and two found a partial compensation (or a net benefit depending on the country).

**Figure 6.**
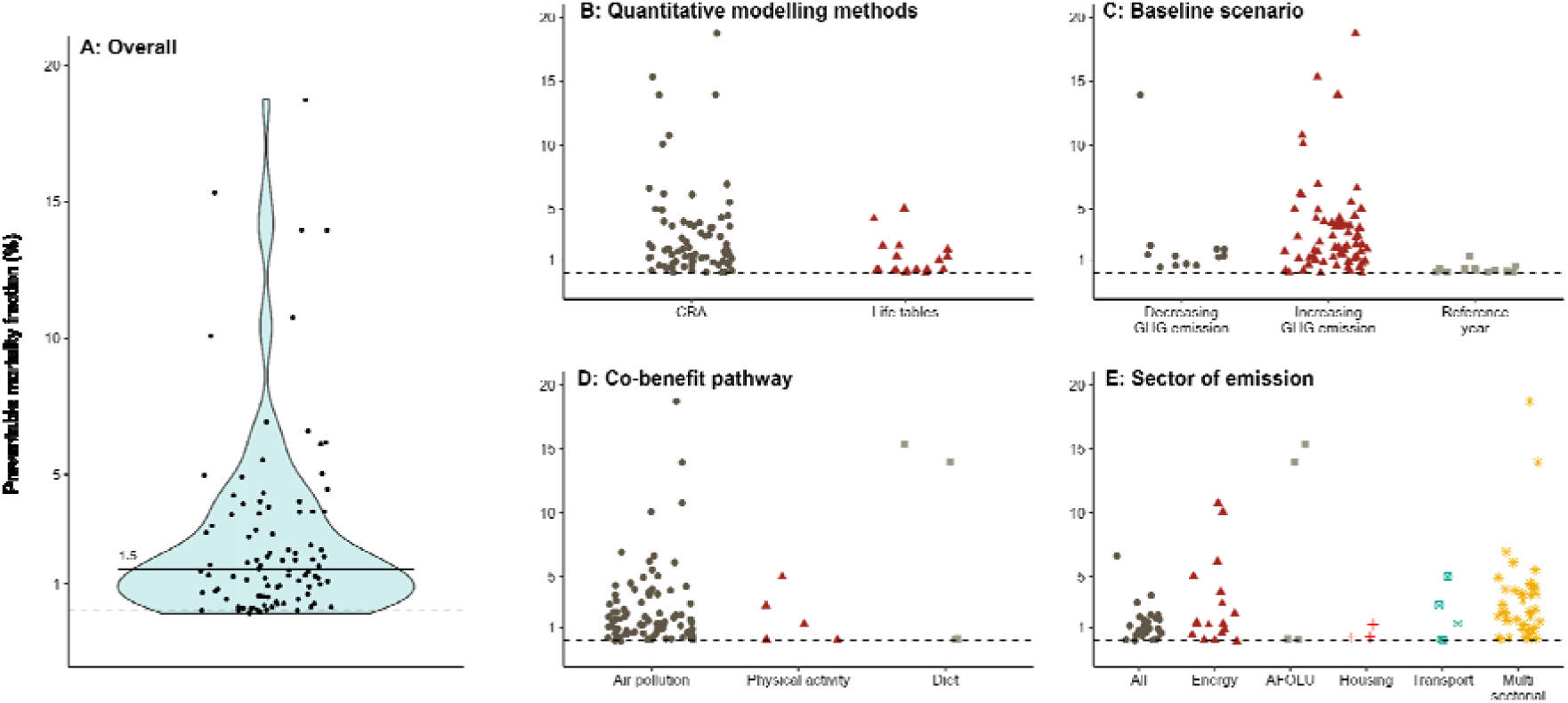
Preventable mortality fraction (%) across net-zero scenarios. We depicted all scalable mortality fractions from our total sample (A) and stratified by health impact assessment methods (B), choice of the baseline scenario (C), type of co-benefit pathway (D) or sector of emission (E). *Horizontal bar represents the median value of preventable mortality (%). CRA: Comparative risk assessment*.

#### Health impact across emission sectors and pathways of co-benefits

Most studies focused only on air pollution in association with one or several emission sectors (Figures 6D and S2), with a wide amplitude of health impacts, as for physical activity and diet pathways.

Regarding the most frequently studied air pollutants, fine particulate matter <2.5µm (PM_2.5_) and ozone (O_3_), the sectors associated with the largest health co-benefits were industry, household, energy, transport and agriculture.^26,42,46,68^ Population density, the sectors of emissions and baseline levels represented important drivers of potential health benefits arising through better air quality.^24,26,37,47,66^ Health co-benefits from decreasing air pollution arose mainly from reduced acute and chronic cardiovascular and respiratory tract diseases.^31,32,48,61^

Increased physical activity also generated substantial public health benefits, which were comparable to the gains expected by large scale health prevention interventions.^52^ In many countries, attainment of net-zero emissions yielded larger co-benefits through dietary shifts, compared to air pollution reduction or active travel.^5^ The pathway yielding the greatest health benefits depended on regional context and the number of mitigation actions modelled.^5,6^

#### Health impact across the typology of net-zero scenarios

Due to a higher potential for reducing air pollution, a scenario that implemented demand reduction policies provided greater health benefits than an energy decarbonization scenario.^17^ Greater benefits were expected if the energy sector was based on renewable instead of carbon capture and storage technologies.^31^ “Health in all policies” scenarios (electrification and clean renewable energy) yielded four times more health co-benefits than financial instrument (combustible renewable fuels).^66^ A city-level study (Beijing) found that developing active travel and public transport yielded higher health co-benefits than the electrification of private vehicles (even without accounting for increased physical activity).^57^ Different socio-economic projections, priorities given and levels of ambition yielded very different health impacts,^19^ especially for physical activity and diet.^6^

#### Equity impact and regional disparities in net-zero scenarios

Very few studies explored the distribution of health impacts regarding socially and economically marginalized populations (n=6). In India, health benefits of net-zero emission scenarios were modelled to be greater for men, urban and high socio-demographic index population.^54^ The implementation of integrated climate, air quality, and clean energy access interventions had a synergistic impact, avoiding millions of stunted children, particularly for the most disadvantaged children and geographic regions.^53^

Ambitious GHG reduction efforts in California provided substantial health co-benefits, especially for residents of disadvantaged communities.^64^ In the US, the enhanced electrification of the transport sector was shown to benefit disadvantaged communities more effectively than building electrification.^65^ Accounting for air pollution-related health impacts showed that climate policies have the potential to reduce inequality and increase welfare at several geographical scales, partly because the most disadvantaged communities were more exposed in some regions.^16,67^ However, even if inequalities were reduced with air quality improvements, they would remain high as long as control measures do not target lower-income regions.^20^

Partially due to a high baseline exposure and population density, air pollution co-benefits were the greatest for China (Figure S3) and India.^5,15–17,20–22^ In G20 countries, benefits were mainly attributable_26_ to PM_2.5_ emission reduction. Mitigation policies affecting air pollution emissions had substantial transboundary health impacts, with the transport sector being a major contributor to these benefits.^13,26^ Carbon trading based on historical mitigation rate and low-carbon investment transfer across regions improved the efficiency of global mitigation actions in some contexts.^14^ Disparities in health impacts were also influenced by population aging, which is expected to increase in the coming years. However, the health co-benefits arising from air pollution mitigation have the potential to offset the effects of population ageing, even for a rapidly ageing country such as China.^41,43–45,59^

## DISCUSSION

### Review findings

Studies assessing the health impact of scenarios aiming at net-zero emissions show public health co-benefits arising from a wide array of scenarios, emission sectors, and co-benefit pathways (Figure 4). 98% of scenarios (94 out of 96) found favourable health impacts that depended on the scenario assumptions, co-benefits pathways and region of implementation. Half of scenarios yielded more than 1.5% of preventable mortality fraction. However, the preventable fraction cannot simply be extrapolated from one setting to another because of the heterogeneity in co-benefit pathways, demographic characteristics, modelling methods and assumptions. A large majority of studies that compared implementation costs with monetized health benefits (11 out of 13) reported that the costs of net-zero policies would be offset by the economic gains provided by health benefits.

The available evidence mostly focused on three major health pathways, namely dietary risks, air pollution and physical inactivity, that have been estimated to be responsible for respectively up to 7, 8 and 4 million global deaths annually.^12,69,70^ Similarly to improved dietary patterns, reduced exposure to air pollution would have the potential to yield very important health benefits, especially in high-density and polluted regions.^5^ More comprehensive policies also targeting household air quality could yield larger health benefits in some regions.^6^ Active transport policies also have a great potential where the lack of physical activity already induces a high health burden.^5^

Our review identified several sources of variability in the assessed impacts. In the reviewed studies, most health impacts were assessed either by CRA or lifetable approaches. CRA is a simpler approach but might overestimate health impacts because it completely averts a proportion of deaths. Lifetable approaches adopt a more realistic model of deaths over time, as they account for age-specific mortality in the population.^71^ The assumptions regarding the baseline scenario, especially the evolution of GHG emissions, might affect the magnitude of predicted health outcomes (Figure 6C).

Explored scenarios and settings were also highly variable. Energy decarbonization based on various technologies received the highest attention, while many net-zero scenarios were not explicit in the transformations assumed to achieve net-zero. Despite their high mitigation potential and synergy with well-being, demand reduction strategies were often marginalized in climate policy and scenarios (Figure S1), with many studies failing to specify implementation mechanisms.^4,72^ A majority of studies were performed in high-income regions (Figure S4) and only a few addressed health inequalities despite their relevance for public health and environmental justice.^73^

### Implication of the results

Given the long residence time of some GHGs (especially CO_2_) in the atmosphere, accelerated and equitable mitigation actions have the potential to attain net-zero emissions only at mid-to long-term, depending on the emission sector (2030-35 for AFOLU and 2050 for the industry).^3^ Conversely, these same actions have the potential to improve health and well-being in the near term^2^ by improving cardio-vascular, respiratory and mental health outcomes associated with co-benefits pathways ^74,75^ particularly from air pollution, diet and physical activity.^6^

Another important feature of health co-benefits of climate mitigation policies highlighted by this review is their largely unconditional nature. From a climate perspective, mitigation actions require to be implemented in a large part of countries and regions to allow for a control of global warming. This nature of climate benefits, which are conditional to global coordinated actions, may be prone to the free-rider problem, where actors do not actively contribute to efforts while expecting to take advantage from collective benefits. Conversely, most of the studies projecting net-zero scenarios reported important health co-benefits while making no specific assumption regarding global coordinated climate actions. In other words, health co-benefits of mitigation policies are largely unconditioned to climate action from other countries or regions, and therefore likely less affected by the free-rider problem. For some pathways (such as physical activity and diet), the health benefits are restricted to the territories that implement the policies. For air quality, the magnitude of health benefits partially depends on the policies implemented by neighbouring countries,^13,26^ but out of the 35 studies assessing air pollution pathway at a national or sub-national scale, 34 revealed that net-zero policies would bring significant local air quality benefits, independently of the actions taken in neighbouring countries.

Relying on monetary valuation of health impacts, studies have shown that health co-benefits of climate policies have the potential to outweigh the costs of net-zero policies, depending on the region, with India and China showing the largest benefits. The Intergovernmental Panel on Climate Change (IPCC) also reports that the global benefits of climate policies (not accounting for health) exceed the cost of mitigation.^2^ Economic impact assessments anticipate other benefits directly or indirectly affecting human health, such as the net creation of millions of jobs, fewer work loss days and tens of billions of dollars for labour productivity, crop yield increase, reduced hospital expenditures ^13,25,55,67^ and a more resilient energy system.^18^

### Research gaps

The high heterogeneity of retrieved studies regarding scenarios, emission sectors, co-benefit pathways and modelling approaches prevented us from drawing conclusions about a clear ranking of co-benefits pathways in terms of potential health impact. In addition, our comparison of health impacts does not account for factors that could potentially lead to differences across studies, particularly due to variations in locations and study populations.

While our review highlighted important health and economic benefits, numerous health impacts remain underestimated. For instance, modal shift to active mode of transportation could provide additional health co-benefits by reducing noise exposure.^76^ Included HIAs also fail to address mental health impacts, despite evidences suggesting an association between air quality and physical activity with mental health.^74,77^ Adaptation measures not accounted for, such as urban green space, also have the potential to yield substantial health benefits.^78^ Incorporating household pollution is essential for assessing potentially detrimental health impacts associated with poorly ventilated housing.^63^ Lastly, only one study considered the impact of prenatal environmental exposures.^53^

Uncertainties in health impact quantification also result from difficulties in considering multiple parameters such as specific exposure-response functions (across age, sex or social factors) or the specific distribution of exposures among the studied population. For each mitigation action, there are also potential positive synergistic effects that can be hard to account for in quantitative assessments, such as reduced air pollution emissions along with changes in active transport and dietary patterns. Conversely, extreme climate hazards can restrain cycling behaviours, and health impacts from combined air pollution and heat exposure are exacerbated.^79^ Prospective assessments also assume a consistent healthcare system efficiency across all scenarios while higher air pollution and temperature are associated with increased hospital admissions.^80^

Many of the studies and scenarios are from high and upper-middle-income regions, where the mitigation efforts are expected to be the greatest, and therefore related societal changes are expected to be important. Whether the magnitude of health co-benefits would be of the same scale in low-income countries remains unclear and will greatly depend on levels of fossil fuel related air pollution, dietary patterns and levels of physical activity.^69^ For instance, evidence suggests that air pollution reduction (and notably household pollution from cooking stoves) could have a high health co-benefit potential in India.^53,54^ Conversely, one study showed that only modest benefits may be expected in Nigeria from sustainable diet policies.^5^

Evidence on the feasibility and acceptability of implementing assessed actions is limited. However, known effective interventions include dietary modifications through education, persuasion, and environmental restructuring.^81^ In the transport sector, active mobility policies are most effective when integrating safe walking and cycling infrastructure with strong public transport support and educational programs. ^82^

Finally, we did not investigate grey literature due to methodological issues, and may thus, for instance, have missed assessments published as reports.

### Perspectives and future directions

Several recommendations for future HIA of net-zero scenarios may be inferred from our review.

First, studies should clearly state and justify which mitigation lever(s) are implied by the policy assessed to better estimate the impacts of diverse type of net-zero emission policies.^17,57,62^ While they gathered a relatively low research interest, demand-side mitigation policies are essential as they have the potential to induce fundamental lifestyle changes that would support the implementation of sustainable and healthy actions.^72^ Policies and actions must extend beyond technological efficiency improvements to address unsustainable systems that drive high energy and material demands, leading to elevated emissions while neglecting healthy environments.^4^ This is particularly evident in the transport sector, where decarbonization policies exclusively focused on technological improvements could exacerbate physical inactivity in the population.^83^

As aging populations can have a significant impact on estimates,^45^ HIAs should prefer lifetable approaches to estimate more accurately health impacts over time while baseline scenarios should include a projection of the studied population to compare the impacts based on the same population pyramid. Prospective HIAs of net-zero scenarios should carefully use adapted vulnerability indicators to assess health impacts when possible and otherwise address inequality impacts qualitatively.^84^ Assessment of energy decarbonization policies should address energy poverty which has environmental justice implications.^85^

The lack of code and data sharing by most of the studies presents a significant barrier to advancing health impact monitoring associated with net-zero scenarios, such as the development of living systematic reviews. Accelerating research and monitoring of health impacts is essential to provide evidence-based and timely feedback to decision-makers.

Finally, our review highlights a need for a standardized framework to assess the health impacts of net-zero emission scenarios. This framework should make use of already existing scalable tools and methods to compare prospective scenarios regarding the evolution of specific exposures, to incorporate a relevant baseline scenario and attribute health impacts across populations over time.

### Conclusion

Our synthesis of the available evidence suggests that achieving net-zero emissions across different sectors would generate large health co-benefits and prevent a considerable fraction of mortality. Therefore, each further delay in implementing transformative changes toward net-zero society may not only increase risks induced by climate change, but also represent a missed opportunity to improve human health. Especially because health co-benefits of climate mitigation policies are expected to manifest in the short term, are not conditioned to global coordinated climate action, and may outweigh the costs of mitigation policies, highlighting these health co-benefits make a strong case for driving impactful mitigation action.

## DECLARATIONS

### Availability of data and materials

All codes, analysis, extraction and quality grid are available in the following GitHub repository: https://github.com/LeoMoutet/revue_syst.

### Authors’ contributions

LM, LT and KJ designed the scope of the review, perfomed the study selection and wrote the original draft of the article. LM and PB extracted the data from included studies. LM perfomed the confidence assessment. RG and JM provided inputs regarding health impact assessment frameworks and health co-benefits. AH and RS contributed to the interpretation of the results. All authors read and approved the final manuscript.

## Data Availability

All codes, analysis, extraction and quality grid are available in the following GitHub repository

https://github.com/LeoMoutet/revue_syst.

## Acknowledgements

The authors would like to thank Audrey De Nazelle for helpful discussions regarding review findings.

## Review protocol

No review protocol was published prior to this study.

