## Supplementary materials for "The public health co-benefits of strategies consistent with net-zero emissions: a systematic review of quantitative studies"

**Table of Contents**

**Supplementary text 1.** Health impact scaling

**Supplementary text 2.** Elaboration of the scenarios

**Table S1.** PRISMA checklist

**Table S2.** Full research equation

**Table S3.** Confidence assessment grid elaboration

**Table S4.** Full description of included studies

**Figure S1.** Typology of net-zero and baseline scenarios

**Figure S2.** Preventable mortality fraction across net-zero scenarios for air pollution exposure, by health impact assessment methods (A) and choice in the baseline scenario (B)

**Figure S3.** Health impact of investigations led in China

**Figure S4.** Countries of 1st authors’ institution

### Supplementary text 1: Health impact scaling

Number of deaths prevented and/or life-years gained were extracted from the included studies. When only life-years gained were estimated, they were converted into deaths prevented in order to compare health impacts across studies.

To do so, we calculated conversion factors from the specific location, age and health outcomes for all-cause mortality reported in the 2021 GBD study (Institute for Health Metrics and Evaluation (IHME), 2024). The conversion factor was linearly extrapolated to the year of interest based on previous years. Mortality projections were based on the number of deaths prevented stated directly in the study or with the GBD tool and the United Nations population prospect 2022, medium fertility variant (<https://population.un.org/wpp/Download/Standard/MostUsed/>). For sub-national studies in California, (Wang et al., 2020; Zhao et al., 2019; Zhu et al., 2022) we retrieved the mortality rate based on the United States Census Bureau, the number of deaths in 2019 given by the GBD and official population projections. (California Department of Finance. Demographic Research Unit., 2023)

In case only cumulative health impacts over a time range were available, we used the average impact for one theoretical year and the population projections of the median year based on the time-scale of the scenario assessed

### Supplementary text 2: Elaboration of the scenarios

- **Comprehensive pathways from a third party** :

(Schmid et al., 2019) = EU reference scenario 2016: Energy, transport and GHG emissions - trends to 2050 ;

(Barban et al., 2022) = NegaWatt scenario on transport demand for France ;

(Milner et al., 2023) = UK Committee on Climate Change, 6th Carbon Budget report ;

(Williams et al., 2018) = UK Committee on Climate Change, 5th Carbon Budget report ;

(Shrubsole et al., 2015) = UK Committee on Climate Change, 4th Carbon Budget report ;

(Hata et al., 2023) = Japan Agency for Natural Resources and Energy, 2022 ;

(T. Ma et al., 2023) = Synthesis Report 2020 on China’s Carbon Neutrality: China’s New Growth Pathway ;

(Shen et al., 2022) = China Energy Interconnection Carbon Neutrality (CEICN) ;

(Xie et al., 2021) = Based on the government’s targets and analysis ;

(Rafaj et al., 2018) = International Energy Agency’s World Energy Outlook 2017 ;

(Conibear et al., 2022) = International Energy Agency’s World Energy Outlook 2018 ;

(Ortiz et al., 2023) = Virginia Clean Economy Act ;

(Lu et al., 2022) = Beijing city master plan (2016–35) ;

(Zyśk et al., 2020) = Energy and emission scenarios of the REFLEX Project ;

- **Based on NDCs** : (Shihui Zhang et al., 2021); (Phillips, 2021); (Cai et al., 2018); (Nawaz et al., 2023b) ; (Polonik et al., 2021)
- **Based on NDCs and adjoins more ambitious objectives** : (Silu Zhang et al., 2021) ; (Qu et al., 2020) ; (Li et al., 2019); (Wang et al., 2022)

(Hamilton et al., 2021) = Based on NDCs, technical projections (IEA) and hypothesis regarding active transport and dietary patterns;

- **Temperature target only :** (McCollum et al., 2013) ; (Rafaj et al., 2013), (Rafaj et al., 2021) ; (Rauner et al., 2020) ; (Vandyck et al., 2018) ; (H. Wang et al., 2023) ; (Sampedro et al., 2021) ; (Dimitrova et al., 2022) ; (Dimitrova et al., 2021) ; (Shindell et al., 2018)
- **SSP/RCP combination (including temperature targets)** : (X. Ma et al., 2023) ; (Reddington et al., 2023) ; (Shindell et al., 2021) ; (Sampedro et al., 2020) ; (Tang et al., 2022) ; (Y. Wang et al., 2023) ; (Liu et al., 2022) ; (Markandya et al., 2018) ; (Reis et al., 2022) ; (Chen et al., 2018)
- **In-house scenarios made by authors** :

(Creutzig et al., 2012) = Determined iteratively by stakeholder workshops and interviews in each of the corresponding cities and by comparison to benchmark policies in other cities.

(Jacobson et al., 2017) = 139 countries reaching full of energy decarbonation with wind, water and sunlight.

(Zyśk et al., 2021) = Public electricity and heat production sector in Poland will reach a CO2 emission reduction of 95% by 2050 compared to the baseline year 1990.

(Zhu et al., 2022) = Decarbonation measures that reduce pollutant emissions (reliance on renewable electricity, high electrification of light-duty vehicles buses, and off-road equipment, reductions in demand for petroleum refining…).

(Yang et al., 2019) = Assumptions of energy conservation and end-of-pipe measures.

(Xing et al., 2020) = Low carbon policies related to energy conservation for both CO2 and air pollution in all sectors included.

(Nawaz et al., 2023a) = Net-zero CO2 emission target year for each G20 countries.

(Luo et al., 2023) = Achieves linearly zero emissions by 2050, 2045 or with 20% less carbon emission for each period of 5 years.

(Lin et al., 2023) = Carbon emission reduction along with forest carbon sink predictions and end-of-pipe control measures.

(Cheng et al., 2023) = China’s CO2 emission and PM2.5 exposure trajectories from 2015 to 2060 and emission peak.

(Chen et al., 2020) = Decarbonation based on current industrial and energy structure transformation and development plan.

(Zhao et al., 2019) = Based on GHG reduction, technology and resource availability and policy trend.

(Wang et al., 2020) = Sector-based CO2e emission reduction to determine the changes in infrastructure and technology over time necessary to meet the GHG emissions target.

(Mo et al., 2022) = Adjoin end-of-pipe control scenarios to the Integrated Policy Model for China (IPAC) model.

### Table S1. PRISMA checklist

*The PRISMA 2020 statement: an updated guideline for reporting systematic reviews (Page et al., 2021).*

| **Section and Topic** | **Item #** | **Checklist item** | **Location where item is reported** |
| --- | --- | --- | --- |
| **TITLE** | | |  |
| Title | 1 | Identify the report as a systematic review. | L. 1-3 |
| **ABSTRACT** | | |  |
| Abstract | 2 | See the PRISMA 2020 for Abstracts checklist. | L. 23-34 |
| **INTRODUCTION** | | |  |
| Rationale | 3 | Describe the rationale for the review in the context of existing knowledge. | L. 54-61 |
| Objectives | 4 | Provide an explicit statement of the objective(s) or question(s) the review addresses. | L. 62-67 |
| **METHODS** | | |  |
| Eligibility criteria | 5 | Specify the inclusion and exclusion criteria for the review and how studies were grouped for the syntheses. | L. 80-91 |
| Information sources | 6 | Specify all databases, registers, websites, organisations, reference lists and other sources searched or consulted to identify studies. Specify the date when each source was last searched or consulted. | L. 73-75 |
| Search strategy | 7 | Present the full search strategies for all databases, registers and websites, including any filters and limits used. | Table S2 |
| Selection process | 8 | Specify the methods used to decide whether a study met the inclusion criteria of the review, including how many reviewers screened each record and each report retrieved, whether they worked independently, and if applicable, details of automation tools used in the process. | L. 78-79 |
| Data collection process | 9 | Specify the methods used to collect data from reports, including how many reviewers collected data from each report, whether they worked independently, any processes for obtaining or confirming data from study investigators, and if applicable, details of automation tools used in the process. | L. 98-99 |
| Data items | 10a | List and define all outcomes for which data were sought. Specify whether all results that were compatible with each outcome domain in each study were sought (e.g. for all measures, time points, analyses), and if not, the methods used to decide which results to collect. | L. 99-105 |
|  | 10b | List and define all other variables for which data were sought (e.g. participant and intervention characteristics, funding sources). Describe any assumptions made about any missing or unclear information. | L. 106-110 |
| Study risk of bias assessment | 11 | Specify the methods used to assess risk of bias in the included studies, including details of the tool(s) used, how many reviewers assessed each study and whether they worked independently, and if applicable, details of automation tools used in the process. | L. 112-116 |
| Effect measures | 12 | Specify for each outcome the effect measure(s) (e.g. risk ratio, mean difference) used in the synthesis or presentation of results. | L. 118-125 |
| Synthesis methods | 13a | Describe the processes used to decide which studies were eligible for each synthesis (e.g. tabulating the study intervention characteristics and comparing against the planned groups for each synthesis (item #5)). | L. 115-125; Supplementary text 1 |
|  | 13b | Describe any methods required to prepare the data for presentation or synthesis, such as handling of missing summary statistics, or data conversions. |  |
|  | 13c | Describe any methods used to tabulate or visually display results of individual studies and syntheses. |  |
|  | 13d | Describe any methods used to synthesize results and provide a rationale for the choice(s). If meta-analysis was performed, describe the model(s), method(s) to identify the presence and extent of statistical heterogeneity, and software package(s) used. |  |
|  | 13e | Describe any methods used to explore possible causes of heterogeneity among study results (e.g. subgroup analysis, meta-regression). | Not applicable |
|  | 13f | Describe any sensitivity analyses conducted to assess robustness of the synthesized results. |  |
| Reporting bias assessment | 14 | Describe any methods used to assess risk of bias due to missing results in a synthesis (arising from reporting biases). | Not applicable |
| Certainty assessment | 15 | Describe any methods used to assess certainty (or confidence) in the body of evidence for an outcome. | Not applicable |
| **RESULTS** | | |  |
| Study selection | 16a | Describe the results of the search and selection process, from the number of records identified in the search to the number of studies included in the review, ideally using a flow diagram. | L. 130-136; Figure 1 |
|  | 16b | Cite studies that might appear to meet the inclusion criteria, but which were excluded, and explain why they were excluded. | L. 132-133; Figure 1 |
| Study characteristics | 17 | Cite each included study and present its characteristics. | L. 137-141;  Table S4 |
| Risk of bias in studies | 18 | Present assessments of risk of bias for each included study. | Table S4 |
| Results of individual studies | 19 | For all outcomes, present, for each study: (a) summary statistics for each group (where appropriate) and (b) an effect estimate and its precision (e.g. confidence/credible interval), ideally using structured tables or plots. | L. 153-204;  Figure 3 |
| Results of syntheses | 20a | For each synthesis, briefly summarise the characteristics and risk of bias among contributing studies. | Table S4 |
|  | 20b | Present results of all statistical syntheses conducted. If meta-analysis was done, present for each the summary estimate and its precision (e.g. confidence/credible interval) and measures of statistical heterogeneity. If comparing groups, describe the direction of the effect. | Not applicable |
|  | 20c | Present results of all investigations of possible causes of heterogeneity among study results. | L. 241-287 |
|  | 20d | Present results of all sensitivity analyses conducted to assess the robustness of the synthesized results. | Not applicable |
| Reporting biases | 21 | Present assessments of risk of bias due to missing results (arising from reporting biases) for each synthesis assessed. | Not applicable |
| Certainty of evidence | 22 | Present assessments of certainty (or confidence) in the body of evidence for each outcome assessed. | L. 206-211 |
| **DISCUSSION** | | |  |
| Discussion | 23a | Provide a general interpretation of the results in the context of other evidence. | L. 290-318 |
|  | 23b | Discuss any limitations of the evidence included in the review. | L. 355-384 |
|  | 23c | Discuss any limitations of the review processes used. | L. 350-354; L. 385-386 |
|  | 23d | Discuss implications of the results for practice, policy, and future research. | L. 320-348; L. 388-412 |
| **OTHER INFORMATION** | | |  |
| Registration and protocol | 24a | Provide registration information for the review, including register name and registration number, or state that the review was not registered. | L. 69-71 |
|  | 24b | Indicate where the review protocol can be accessed, or state that a protocol was not prepared. |  |
|  | 24c | Describe and explain any amendments to information provided at registration or in the protocol. |  |
| Support | 25 | Describe sources of financial or non-financial support for the review, and the role of the funders or sponsors in the review. | L. 429 |
| Competing interests | 26 | Declare any competing interests of review authors. | L. 427 |
| Availability of data, code and other materials | 27 | Report which of the following are publicly available and where they can be found: template data collection forms; data extracted from included studies; data used for all analyses; analytic code; any other materials used in the review. | L. 125; L. 424-425 |

*PRISMA 2020 for Abstracts Checklist*

| **Section and Topic** | **Item #** | **Checklist item** | **Reported (Yes/No)** |
| --- | --- | --- | --- |
| **TITLE** | | |  |
| Title | 1 | Identify the report as a systematic review. | Yes |
| **BACKGROUND** | | |  |
| Objectives | 2 | Provide an explicit statement of the main objective(s) or question(s) the review addresses. | Yes |
| **METHODS** | | |  |
| Eligibility criteria | 3 | Specify the inclusion and exclusion criteria for the review. | No* |
| Information sources | 4 | Specify the information sources (e.g. databases, registers) used to identify studies and the date when each was last searched. | No* |
| Risk of bias | 5 | Specify the methods used to assess risk of bias in the included studies. | No* |
| Synthesis of results | 6 | Specify the methods used to present and synthesise results. | No* |
| **RESULTS** | | |  |
| Included studies | 7 | Give the total number of included studies and participants and summarise relevant characteristics of studies. | Yes |
| Synthesis of results | 8 | Present results for main outcomes, preferably indicating the number of included studies and participants for each. If meta-analysis was done, report the summary estimate and confidence/credible interval. If comparing groups, indicate the direction of the effect (i.e. which group is favoured). | Yes |
| **DISCUSSION** | | |  |
| Limitations of evidence | 9 | Provide a brief summary of the limitations of the evidence included in the review (e.g. study risk of bias, inconsistency and imprecision). | No* |
| Interpretation | 10 | Provide a general interpretation of the results and important implications. | Yes |
| **OTHER** | | |  |
| Funding | 11 | Specify the primary source of funding for the review. | Yes |
| Registration | 12 | Provide the register name and registration number. | Yes |

*Not present in the abstract due to word limit but developed in the text.

*From:*  Page MJ, McKenzie JE, Bossuyt PM, Boutron I, Hoffmann TC, Mulrow CD, et al. The PRISMA 2020 statement: an updated guideline for reporting systematic reviews. BMJ 2021;372:n71. doi: 10.1136/bmj.n71

### Table S2. Full research equation

PubMed:

| **#1** | health*[Title/Abstract] OR mortality[Title/Abstract] OR death*[Title/Abstract] |
| --- | --- |
| **#2** | net zero[Title/Abstract] OR net-zero[Title/Abstract] OR decarboni*[Title/Abstract] OR transition scenario[Title/Abstract] OR carbon neutrality[Title/Abstract] OR paris agreement [Title/Abstract] OR climate change act[Title/Abstract] OR climate change action* [Title/Abstract] OR climate change acts[Title/Abstract] OR climate change target* [Title/Abstract] OR below 2°C [Title/Abstract] OR below 1.5°C [Title/Abstract] OR limited to 2°C[Title/Abstract] OR 2°C scenario[Title/Abstract] OR 2°C trajectory[Title/Abstract] OR limited to 1.5°C[Title/Abstract] OR 1.5°C scenario[Title/Abstract] OR 1.5°C trajectory[Title/Abstract] |
| **#3** | **#1** AND **#2** |
| **#4** | **#3** Filter: **None** |

Web Of Science:

| **#1** | TS=(health* OR mortality OR death*) |
| --- | --- |
| **#2** | TS=("net zero" OR net-zero OR decarboni* OR "transition scenario" OR "carbon neutrality" OR "paris agreement" OR "climate change act" OR "climate change action*" OR "climate change acts" OR "climate change target*" OR "below 2°C" OR "below 1.5°C" OR “limited to 2°C” OR 2°C scenario OR 2°C trajectory OR limited to 1.5°C OR 1.5°C scenario OR 1.5°C trajectory) |
| **#3** | **#1** AND **#2** |
| **#4** | **#3** Filter: **None** |

Scopus:

| **#1** | TITLE-ABS (health* OR mortality OR mortality*) |
| --- | --- |
| **#2** | TITLE-ABS ("net zero" OR net-zero OR decarboni* OR "transition scenario" OR "carbon neutrality" OR "paris agreement" OR "climate change act" OR "climate change action*" OR "climate change acts" OR "climate change target*" OR "below 2°C" OR "below 1.5°C " OR “limited to 2°C” OR 2°C scenario OR 2°C trajectory OR limited to 1.5°C OR 1.5°C scenario OR 1.5°C trajectory) |
| **#3** | **#1** AND **#2** |
| **#4** | **#3** Filter: **None** |

### Table S3. Confidence assessment grid elaboration

| **Criteria extracted from guidelines** (Hess et al., 2020) | **Adapted criteria** |
| --- | --- |
| Describe mitigation policies and scenarios and their relevant sectors (e.g., finance, energy, transportation, housing, industry, food systems including agriculture). | Describe mitigation policies and baseline scenarios, detailing relevant sectors of emissions, geographic scale and characteristics of the populations. |
| Specify the geographic scale (international, regional, national, subnational, city) and geographic area of interest. |  |
| Describe populations (size and other characteristics) for model baseline and counterfactual scenarios. |  |
| List and describe counterfactual scenarios, ensuring that these reflect current realities in the absence of strong mitigation policies. As appropriate, list and describe other mitigation policies used in the model. |  |
| If applicable, account for equity and describe socially and economically marginalized populations. | Address equity by describing socially and economically marginalized populations, and evaluate the equity impacts of policy adoption |
| Describe equity impacts of policy uptake. |  |
| If applicable, account for and describe populations that are most likely to experience adverse consequences or benefit the most. | If applicable, account for and describe populations that are most likely to experience adverse consequences or benefit the most. |
| Describe how the analyses will account for projected demographic changes | Describe how the analyses will incorporate projected demographic changes and detail the methodology for assigning exposure to mitigation actions, including the proportion of the population exposed over time based on implementation. |
| Describe how exposure(s) to the mitigation action(s) and related downstream exposure(s) is assigned, including, as appropriate, the proportion of the population that is exposed over time as a function of implementation. |  |
| Describe any potential correspondence between mitigation and counterfactual scenarios with SSPs, RCPs, and/or other globally agreed-upon scenarios, either qualitatively through narrative linkages (e.g., if the mitigation and counterfactual scenarios are characteristic of particular scenarios) or quantitatively (e.g., using specific combinations of RCPs and SSPs with numerical correlates for emissions, demographic shifts, etc.). | Describe any potential correspondence between mitigation and counterfactual scenarios with SSPs, RCPs, and/or other globally agreed-upon scenarios, either qualitatively through narrative linkages (e.g., if the mitigation and counterfactual scenarios are characteristic of particular scenarios) or quantitatively (e.g., using specific combinations of RCPs and SSPs with numerical correlates for emissions, demographic shifts, etc.). |
| List and describe data sources used for population and demographic projections. | Provide a comprehensive overview of data sources utilized, including those for population, demographic projections and counterfactual scenarios. |
| Describe data sources used for counterfactual scenarios (for example, emission scenarios). |  |
| Describe and justify sources used for baseline health estimates and demographics (e.g., national vital statistics, the Global Burden of Disease study, etc.). |  |
| Specify and justify the baseline year for data sources used in the model. | Specify the baseline year for data sources used in the model and define time frame and projected time horizons |
| Clearly define and justify time frames and projected time horizons. |  |
| Describe and justify exposure–response functions used. | Describe exposure-response functions used and health response studies used for modeling |
| Describe health response studies used for modeling (e.g., description of sample size, location, timeframe) and justify if health responses were not obtained from systematic reviews or meta-analyses (e.g., use of local studies). |  |
| Define and justify metric(s) for measuring the health of populations (e.g., health metrics such as DALYs, years of life lost (YLL), years lived with disability (YLD), mortality, LLE, hospitalizations, emergency department visits, among others) appropriate for the specified causal pathways and target audience/decision makers. | Define and justify metric(s) for measuring the health of populations (e.g., health metrics such as DALYs, years of life lost (YLL), years lived with disability (YLD), mortality, LLE, hospitalizations, emergency department visits, among others) appropriate for the specified causal pathways and target audience/decision makers. |
| Conduct a sensitivity analysis and describe methods used. | Conduct a sensitivity analysis and describe methods used. |
| Describe general study limitations qualitatively (e.g., describe potential sources of bias and likely impact on findings) and quantitatively (e.g., describe limitations in valid parameterization of model). | Discuss general study limitations and uncertainty qualitatively (e.g., describe potential sources of bias and likely impact on findings) and quantitatively (e.g., describe limitations in valid parameterization of model). |
| Discuss sources of uncertainty that could not be addressed quantitatively. |  |
| Discuss adverse consequences of mitigation actions (potential, hypothetical, or those observed in results) either qualitatively or quantitatively. | Discuss adverse consequences of mitigation actions (potential, hypothetical, or those observed in results) either qualitatively or quantitatively. |
| When legally and ethically possible, openly and publicly share data and code used for HEM studies to facilitate model validation of models by external researchers, collaboration between investigators, and the production of meta-analyses (including through retrospective harmonization), and new studies. If there are legal or ethical limitations to data sharing, these should be explicitly stated. | When legally and ethically possible, openly and publicly share data and code used for HEM studies to facilitate model validation of models by external researchers, collaboration between investigators, and the production of meta-analyses (including through retrospective harmonization), and new studies. If there are legal or ethical limitations to data sharing, these should be explicitly stated. |

### Table S4. Full description of included studies

*CCS: Carbon capture and storage*

| **Author, date** | **Region & Time scale** | **Modelling :**  **1. Exposure**  **2. Health**  **3. Economic** | **Sector of emission** | **- Health outcome**  **- Age range** | **Category of net-zero scenario(s)** | **Confidence assessment score** |
| --- | --- | --- | --- | --- | --- | --- |
| **Air pollution** | | | | | |  |
| (Cai et al., 2018) | China,  2010-2050 | 1. WRF/ CMAQ  2. Lifetables (IHIA)  3. VSL | Energy | - Mortality  - All | Not detailed : NDCs | Yes: 8  Yes partially: 2  Unclear: 0  No: 3 |
| (Chen et al., 2020) | China,  2015-2050 | 1. LEAP  2. CRA (IF)  3. CCE | All-encompassing | - Mortality/ Morbidity  - All | Energy decarbonation (renewable & CCS) | Yes: 7  Yes partially: 1  Unclear: 0  No: 5 |
| (Chen et al., 2018) | China,  2015-2055 | 1. CMIP5  2. CRA | All-encompassing | - Mortality  - All | Not detailed : RCP | Yes: 1  Yes partially: 9  Unclear: 0  No: 3 |
| (Cheng et al., 2023) | China,  2020-2060 | 1. WRF/ CMAQ / GCAM / DPEC  2. CRA | Energy, industry, transport, housing | - Mortality  - All | Energy decarbonation (CCS) | Yes: 10  Yes partially: 1  Unclear: 0  No: 2 |
| (Conibear et al., 2022) | China,  2020-2050 | 1. WRF/ ECLIPSE/  GAINS  2. CRA (PAF/GEMM) | Residential, industry, transport, agriculture, energy | - Mortality  - 25+ | Not detailed: SSP-RCP | Yes: 7  Yes partially: 2  Unclear: 0  No: 4 |
| (Dimitrova et al., 2021) | India,  2010-2050 | 1. GAINS  2. CRA (GEMM) | Energy, transport, industry, agriculture | - Mortality/ Life expectancy  - All | Not detailed: NDC & Target temperature | Yes: 9  Yes partially: 1  Unclear: 0  No: 3 |
| (Hata et al., 2023) | Japan,  2015-2050 | 1. AIM/ GAINS/ CMAQ  2. Lifetables (BenMap) | All-encompassing | - Mortality  - All | Energy decarbonation  (Electrification & hydrogen) | Yes: 5  Yes partially: 2  Unclear: 1  No: 5 |
|  |  |  |  |  | Sufficiency |  |
| (Li et al., 2019) | China,  2015-2050 | 1. TIMES/ GAINS  2. CRA | Energy, industry, transport, housing | - Mortality  - All | Not detailed: NDC & Target temperature | Yes: 7  Yes partially: 2  Unclear: 0  No: 4 |
| (Lin et al., 2023) | Shaanxi (China),  2017-2060 | 1. CGE/ GAINS  2. CRA  3. VSL + COI | Service, transport, construction, industry, energy | - Mortality/ Morbidity  - All | Energy decarbonation (carbon intensity & CCS) | Yes: 6  Yes partially: 1  Unclear: 0  No: 6 |
| (Liu et al., 2022) | China,  2015-2050 | 1. GCAM/ DPEC/ WRF/ CMAQ  2. CRA | Energy, industry, housing, transport, food system, solvent | - Mortality  - 25+ | Energy decarbonation (CCS) | Yes: 11  Yes partially:  Unclear: 0  No: 2 |
| (Lu et al., 2022) | Beijing (China),  2020-2050 | 1. GAINS  2. CRA (GEMM)  3. VSL + Social cost of carbon | Transport | - Mortality  - 25+ | Energy decarbonation (Electrification)/ | Yes: 10  Yes partially: 1  Unclear: 0  No: 2 |
|  |  |  |  |  | Health in climate policies |  |
| (Luo et al., 2023) | China,  2022-2050 | 1. GridPath  2. CRA (InMAP-China)  3. VSL | Energy | - None  - All | Energy decarbonation (Renewable, CCS, nuclear) | Yes: 5  Yes partially: 3  Unclear: 1  No: 4 |
| (X. Ma et al., 2023) | Shandong (China),  2017-2060 | 1. GCAM/ DPEC/ MEIC  2. CRA | Energy, industry, housing, transport, food system, solvent | - Acute myocardial infarction  - 18+ | Energy decarbonation (CCS) | Yes: 10  Yes partially: 0  Unclear: 1  No: 2 |
| (T. Ma et al., 2023) | China,  2014-2060 | 1. IMED\|TEC/ GAINS  2. CRA (GEMM & IMED\|HEL) | Energy | - Mortality  - All | Energy decarbonation (Electrification) | Yes: 8  Yes partially: 2  Unclear: 0  No: 3 |
| (Markandya et al., 2018) | World,  2020-2050 | 1. GCAM  2. CRA (TM5/ FASST)  3. VSL | All-encompassing | - Mortality  - All | Not detailed (Target temperature) | Yes: 10  Yes partially: 1  Unclear: 0  No: 3 |
| (McCollum et al., 2013) | World,  2010-2030 | 1. MESSAGE/ GAINS/ TMS  2. CRA (TM5) | Energy | - DALYs  - 30+ | Health in climate policies | Yes: 6  Yes partially: 2  Unclear: 0  No: 5 |
| (Mo et al., 2022) | Guangdong (China),  2015-2050 | 1. WRF/ MOSAIC  2. CRA | Energy | - Mortality  - 25+ | Not detailed | Yes: 7  Yes partially: 2  Unclear: 0  No: 4 |
| (Nawaz et al., 2023a) | 41 countries,  2010-2040 | 1. GEOSChem  2. CRA | Agriculture, energy, industry, housing, shipping, transport, waste | - Mortality  - All | Not detailed | Yes: 8  Yes partially: 2  Unclear: 1  No: 2 |
| (Nawaz et al., 2023b) | Santiago (Chile),  2016-2050 | 1. GEOSChem  2. CRA | Energy, industry, transport, agriculture, housing | - Mortality  - All | Health in climate policies | Yes: 8  Yes partially: 2  Unclear: 0  No: 3 |
| (Ortiz et al., 2023) | Virginia (USA),  2016-2045 | 1. "Virginia Clean Economy Act  2. CRA | Energy | - Mortality  - 20+ | Energy decarbonation (Renewable) | Yes: 9  Yes partially: 1  Unclear: 0  No: 3 |
| (Phillips, 2021) | South Korea,  2022-2050 | 1. National projections  2. Lifetables (AirQ+) | Energy, industry, transport, agriculture, waste, building | - Life-years  - 30+ | Not detailed: NDC | Yes: 7  Yes partially: 3  Unclear: 0  No: 3 |
| (Polonik et al., 2021) | World,  2018-2030 | 1. CMIP6 GISS-E2.1  2. CRA | Agriculture, energy, industry, transportation, residential/ commercial, solvents, waste, and shipping | - Mortality  - All | Health in climate policies | Yes: 8  Yes partially: 2  Unclear: 0  No: 3 |
| (Qu et al., 2020) | China,  2015-2030 | 1. CREM/ CMAQ  2. CRA (CREM-HE)  3. VSL | Agriculture, energy, industry, transport, services | - Mortality/ Morbidity  - All | Not detailed: NDC & Target temperature | Yes: 6  Yes partially: 2  Unclear: 0  No: 5 |
| (Rafaj et al., 2013) | EU-27, China, India,  2005-2050 | 1. POLES/ GAINS  2. Lifetables (GAINS) | Energy, industry, transport, housing | - Mortality/ Life expectancy  - All | Not detailed: Global CO2 reduction | Yes: 6  Yes partially: 1  Unclear: 0  No: 6 |
| (Rafaj et al., 2021) | 16 Asian country, 2015-2050 | 1. GAINS  2. CRA | Energy, industry, transport | - Mortality  - All | Not detailed: NDC & Target temperature | Yes: 9  Yes partially: 2  Unclear: 0  No: 2 |
| (Rauner et al., 2020) | World,  2015-2050 | 1. REMIND/ GAINS  2. CRA (TM5-FASST)  3. VSL | All-encompassing | - Mortality  - All | Not detailed: NDC & Target temperature | Yes: 8  Yes partially: 2  Unclear: 0  No: 3 |
| (Reddington et al., 2023) | World,  2020-2100 | 1. CMIP6  2. CRA (GEMM) | All-encompassing | - Mortality  - 25+ | Not detailed | Yes: 8  Yes partially: 2  Unclear: 0  No: 3 |
| (Reis et al., 2022) | World,  2015-2050 | 1. WITCH/ GAINS  2. CRA (FASST) | All-encompassing | - Mortality  - All | Financial instrument | Yes: 8  Yes partially: 2  Unclear: 0  No: 3 |
| (Sampedro et al., 2020) | World,  2020-2050 | 1. GAINS/ TM5/ FASST  2. CRA (FASST)  3. VSL | Energy | - Mortality  - 30+ (IHD, stroke, COPD, LC)  0-5 (ALRI) | Energy decarbonation (CCS & Nuclear) | Yes: 10  Yes partially: 1  Unclear: 0  No: 2 |
|  |  |  |  |  | Sufficiency |  |
| (Sampedro et al., 2021) | World,  2020-2050 | 1. GAINS/ TM5/ FASST  2. CRA (FASST) | Energy, industry, housing, transport, shipping, land use, waste | - Mortality  - 30+ (IHD, stroke, COPD, LC)  0-5 (ALRI) | Health in climate policies | Yes: 9  Yes partially: 2  Unclear: 0  No: 2 |
| (Schmid et al., 2019) | EU-28,  2015-2050 | 1. TIME PanEU  2. CRA (EcoSense)  3. VSL (EcoSense) | Energy | - None  - All | Energy decarbonation (renewable) | Yes: 5  Yes partially: 2  Unclear: 1  No: 5 |
|  |  |  |  |  | Health in climate policies |  |
| (Shen et al., 2022) | China,  2020-2060 | 1. MESSAGE/ GLOBIOM/ GAINS  2. CRA  3. VSL | Industry, transport, housing | - Mortality  - All | Energy decarbonation | Yes: 6  Yes partially: 2  Unclear: 0  No: 5 |
| (Shindell et al., 2018) | World, 2020-2100 | 1. GISS  2. CRA | All-encompassing | - Mortality  - All | Not detailed: Target temperature | Yes: 8  Yes partially: 2  Unclear: 0  No: 3 |
| (Shindell et al., 2021) | China,  2020-2060 | 1. GISS  2. CRA  3. VSL | All-encompassing | - Mortality  - 30+ | Not detailed: SSP-RCP | Yes: 7  Yes partially: 1  Unclear: 0  No: 5 |
| (Tang et al., 2022) | China,  2010-2050 | 1. WRFChem/ GAINS  2. CRA (GEMM)  3. VSL | All-encompassing | - Mortality  - 25+ | Not detailed: SSP-RCP | Yes: 10  Yes partially: 1  Unclear: 0  No: 2 |
| (Vandyck et al., 2018) | World,  2020-2050 | 1. POLES-JRC/ ECLIPSE/ GAINS/ TM5/ FASST  2. CRA | All-encompassing | - Mortality  - All | Energy decarbonation | Yes: 6  Yes partially: 2  Unclear: 0  No: 5 |
| (Wang et al., 2020) | California (USA),  2010-2050 | 1. WRF-Chem/ MEET-CA  2. Lifetables (BenMap)  3. VSL | Energy | - Mortality  - All | Energy decarbonation (renewable) | Yes: 10  Yes partially: 2  Unclear: 0  No: 1 |
| (Wang et al., 2022) | World,  2015-2050 | 1. GAINS/ CAM-Chem  2. CRA (IMED\|HEL)  3. VSL | All-encompassing | - Mortality  - All | Not detailed: NDC & Target temperature | Yes: 8  Yes partially: 2  Unclear: 0  No: 3 |
| (H. Wang et al., 2023) | World,  2100-2020 | 1. GTIME/ GAINS  2. CRA (TM5/ FASST)  3. VSL | All-encompassing | - Mortality  - 30+ (IHD, cerebro-vascular, COPD, LC)  5+ (ALRI) | Not detailed: Target temperature | Yes: 6  Yes partially: 2  Unclear: 1  No: 4 |
| (Y. Wang et al., 2023) | China,  2015-2060 | 1. GEOSChem  2. CRA (GEMM) | Agriculture, energy, industry, transport, residential, commercial, solvent, waste, shipping, natural burning | - Mortality  - 25+ | Not detailed | Yes: 7  Yes partially: 2  Unclear: 0  No: 4 |
| (Williams et al., 2018) | UK,  2011-2154 | 1. UK-TIMES/ ECLIPSE/ WRF/ CMAQ  2. Lifetables | Energy | - Life-years  - All | Energy decarbonation (Nuclear) | Yes: 8  Yes partially: 1  Unclear: 0  No: 4 |
| (Xie et al., 2021) | Anhui (China),  2020-2060 | 1. Emission factor  2. CRA  3. Unit value of health outcomes | Energy | - Mortality/ Morbidity  - All | Energy decarbonation (Electrification) | Yes: 8  Yes partially: 0  Unclear: 0  No: 5 |
|  |  |  |  |  | Sufficiency |  |
| (Xing et al., 2020) | China,  2015-2035 | 1. GCAM/ CMAQ  2. CRA  3. VSL (BenMap-CE) | Housing, industry, transport, energy | - Mortality  - All | Health in climate policies | Yes: 6  Yes partially: 3  Unclear: 0  No: 4 |
| (Yang et al., 2019) | China,  2010-2050 | 1. China-MAPLE/ inF  2. CRA  3. VSL | Energy | - Mortality  - All | Energy decarbonation (CCS) | Yes: 7  Yes partially: 2  Unclear: 0  No: 4 |
|  |  |  |  |  | Sufficiency |  |
| (Silu Zhang et al., 2021) | Sichuan (China),  2015-2035 | 1. IMEG\|CGE/ GAINS  2. CRA (IMED\|HEL/ GEMM)  3. VSL + COI | Energy | - Mortality  - All | Energy decarbonation (CCS) | Yes: 7  Yes partially: 1  Unclear: 0  No: 5 |
| (Shihui Zhang et al., 2021) | China,  2020-2060 | 1. CHEER-LCT/AIR  2. CRA (CHEER-HA)  3. VSL + COI | Energy | - Mortality/ Life expectancy  - All | Energy decarbonation (CCS & renewable) | Yes: 8  Yes partially: 0  Unclear: 0  No: 5 |
| (Zhao et al., 2019) | California (USA),  2010-2050 | 1. WRF-Chem/ CMAQ  2. Lifetables (BenMap-CE) | Energy, transport, residential, commercial, industry, agriculture | - Mortality  - All | Health in climate policies | Yes: 5  Yes partially: 1  Unclear: 0  No: 7 |
|  |  |  |  |  | Financial instrument |  |
| (Zhu et al., 2022) | California (USA),  2018-2050 | 1. WRF-Chem/ SMOKE/ CMAQ  2. Lifetables (BenMap-CE) | Transport, housing | - Mortality  - All | Energy decarbonation (electrification) | Yes: 9  Yes partially: 0  Unclear: 0  No: 4 |
| (Zyśk et al., 2020) | Poland,  2015-2050 | 1. Polyphemus  2. Life tables (πESA)  3. Unit damage costs | Energy | - Life-years / Morbidity  - All | Energy decarbonation (renewable) | Yes: 6  Yes partially: 1  Unclear: 0  No: 6 |
| (Zyśk et al., 2021) | Poland,  2018-2050 | 1. Polyphemus  2. Life tables  3. Unit damage costs | Energy | - None  - All | Energy decarbonation (nuclear & renewable) | Yes: 6  Yes partially: 0  Unclear: 0  No: 7 |
| **Air and indoor pollution** | | | | | |  |
| (Dimitrova et al., 2022) | India,  2010-2050 | 1. GAINS/ MESSAGE-GLOBIOM  2. Microsimulation | All-encompassing | - Child stunting  - 0-5 | Health in climate policies | Yes: 11  Yes partially: 1  Unclear: 1  No: 0 |
| (Jacobson et al., 2017) | 139 countries | 1. GATOR-GCMOM  2. CRA  3. VSL + morbidity | Energy | - Mortality | Energy decarbonation (renewable) | Yes: 3  Yes partially: 1  Unclear: 3  No: 6 |
| (Rafaj et al., 2018) | China, India, EU, Indonesia, South Africa, 2015-2040 | 1. WEM_GAINS  2. CRA | Energy, industry, transport, housing, agriculture, waste | - Mortality  - All | Health in climate policies | Yes: 6  Yes partially: 2  Unclear: 0  No: 5 |
| **Indoor pollution** | | | | | |  |
| (Shrubsole et al., 2015) | London, Milton Keynes (UK), 2010-2050 | 1. SCRIBE  2. Lifetables (IOMLIFET) | Housing | - Life-years  - 40+ | Energy decarbonation (electrification) | Yes: 7  Yes partially: 2  Unclear: 0  No: 4 |
| **Physical activity** | | | | | |  |
| (Barban et al., 2022) | France,  2021-2050 | 1. Negawatt  2. Lifetables  3. VSL | Transport | - Mortality/  Life expectancy/  Life-years  - 20-84 | Sufficiency | Yes: 12  Yes partially: 0  Unclear: 0  No: 1 |
| (Creutzig et al., 2012) | Barcelona, Freiburg, Malmö, Sofia,  2010-2040 | 1. Theoretical policies  2. CRA (HEAT) | Transport | - Mortality  - All | Health in climate policies | Yes: 7  Yes partially: 1  Unclear: 0  No: 5 |
|  |  |  |  |  | Financial instrument |  |
| **Pollution, physical activity and diet** | | | | | |  |
| (Hamilton et al., 2021) | Brazil, China, Germany, India, Indonesia, Nigeria, South Africa, UK, USA,  2015-2040 | 1. GAINS (pollution) / IMPACT (diet)/ Theoretical policies (physical activity)  2. CRA | Energy/  Food system/  Transport | - Mortality  - 0-85 (air pollution) | Health in climate policies | Yes: 8  Yes partially: 2  Unclear: 0  No: 3 |
| (Milner et al., 2023) | England, Wales,  2021-2100 | 1. CRAFT/ SCRIBE (pollution)  ITHIM (physical activity)  Theoretical policies (Diet)  2. Lifetables | Energy/  Food system/  Transport/ Housing | - Life-years  - 25+ (air pollution)/ 15+ (physical activity) | Energy decarbonation | Yes: 9  Yes partially: 2  Unclear: 0  No: 2 |
|  |  |  |  |  | Sufficiency |  |

### Figure S1. Typology of net-zero and baseline scenarios

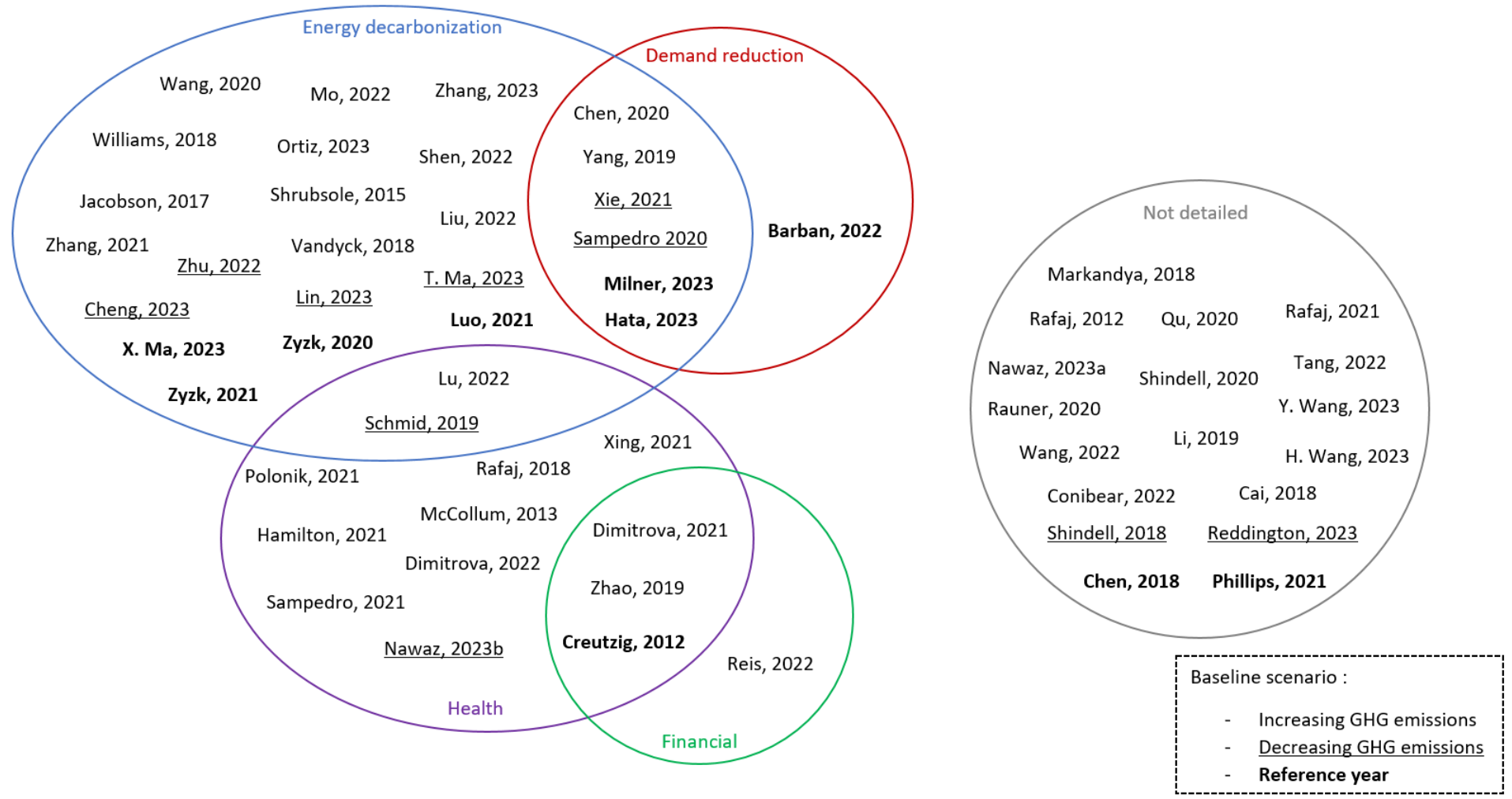

Example of actions for each category:

| **Typology** | **Example of article** | **Example of action** | **Quote from article** |
| --- | --- | --- | --- |
| Energy decarbonization | (Ortiz et al., 2023) | Electrification | « *… transitioning to fossil fuel-free electric generation…* » |
| Demand reduction | (Barban et al., 2022) | Demand-side mitigation | *« …the scenario assumes an overall decrease in transportation demand… »* |
| Health in climate policy | (Dimitrova et al., 2022) | Air quality target | « *…mitigation efforts with targeted air quality and energy access policies…* » |
| Financial instrument | (Reis et al., 2022) | Carbon pricing | « …*model the achieved TTs by imposing a global uniform carbon tax… »* |

TTs: Temperature Targets

### Figure S2. Preventable mortality fraction across net-zero scenarios for air pollution exposure, by health impact assessment methods (A) and choice in the baseline scenario (B)

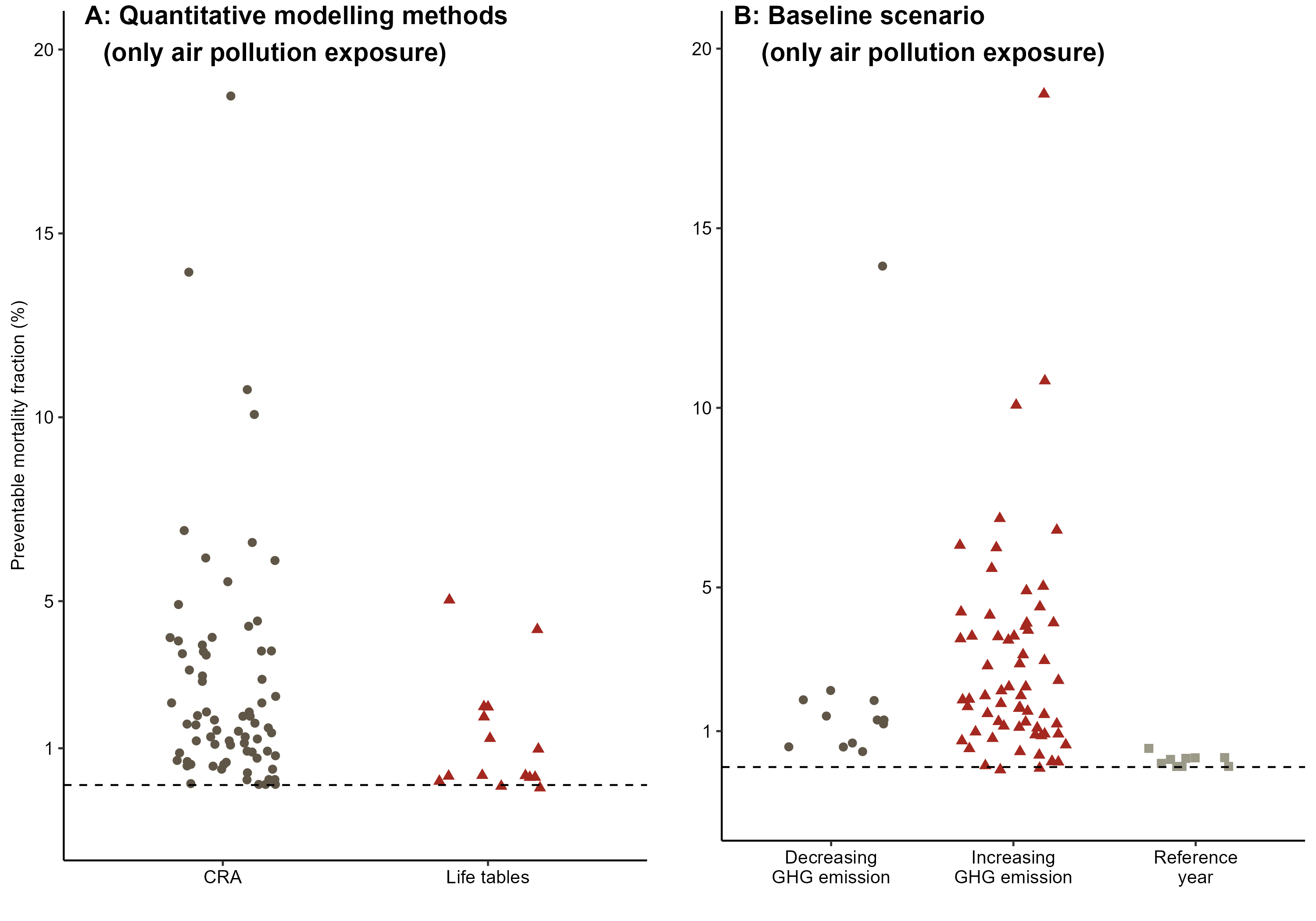

### Figure S3. Health impact of investigations led in China

*Horizontal bar represents the median value of preventable mortality (%)*

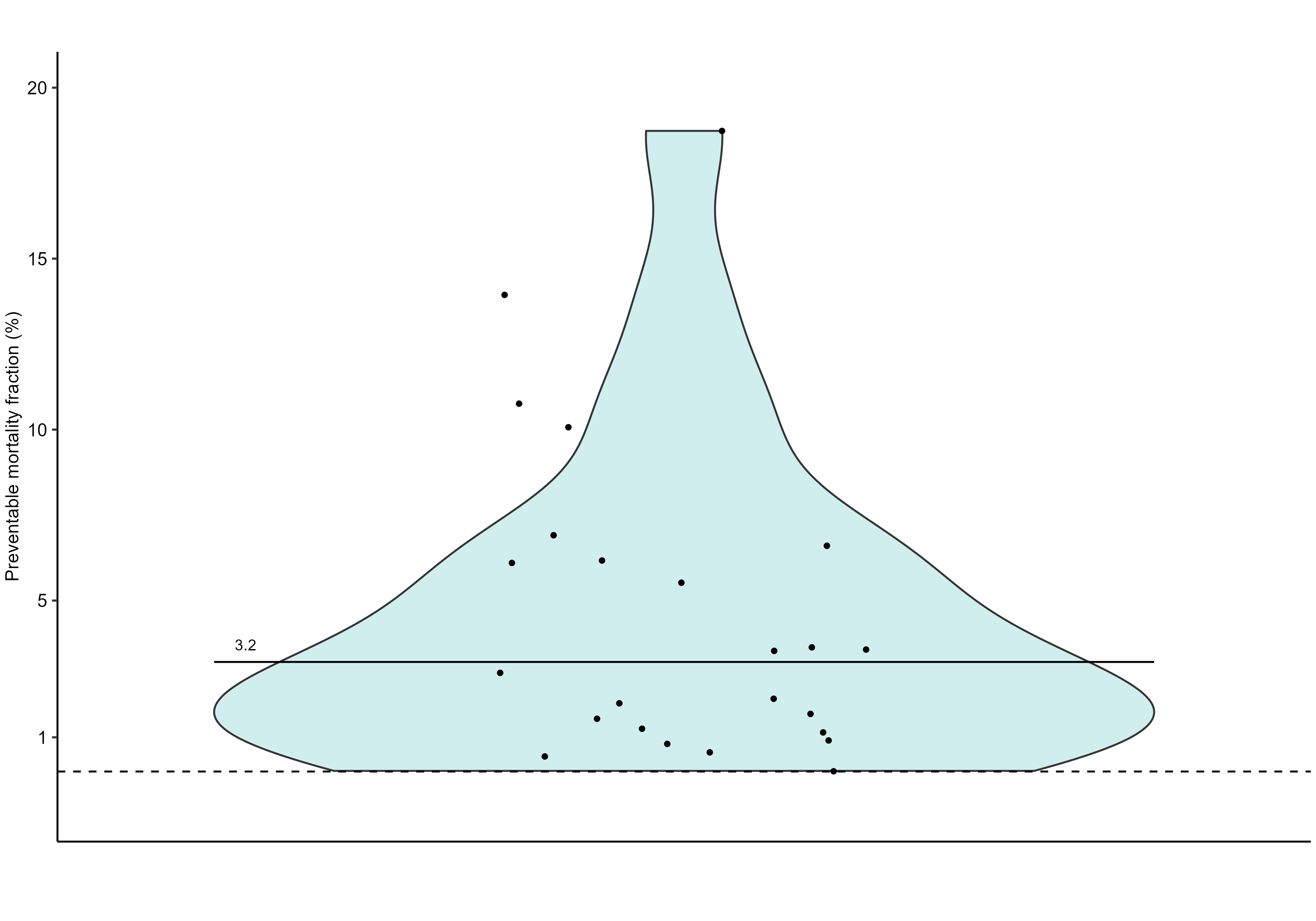

### Figure S4. Countries of 1st authors’ institution

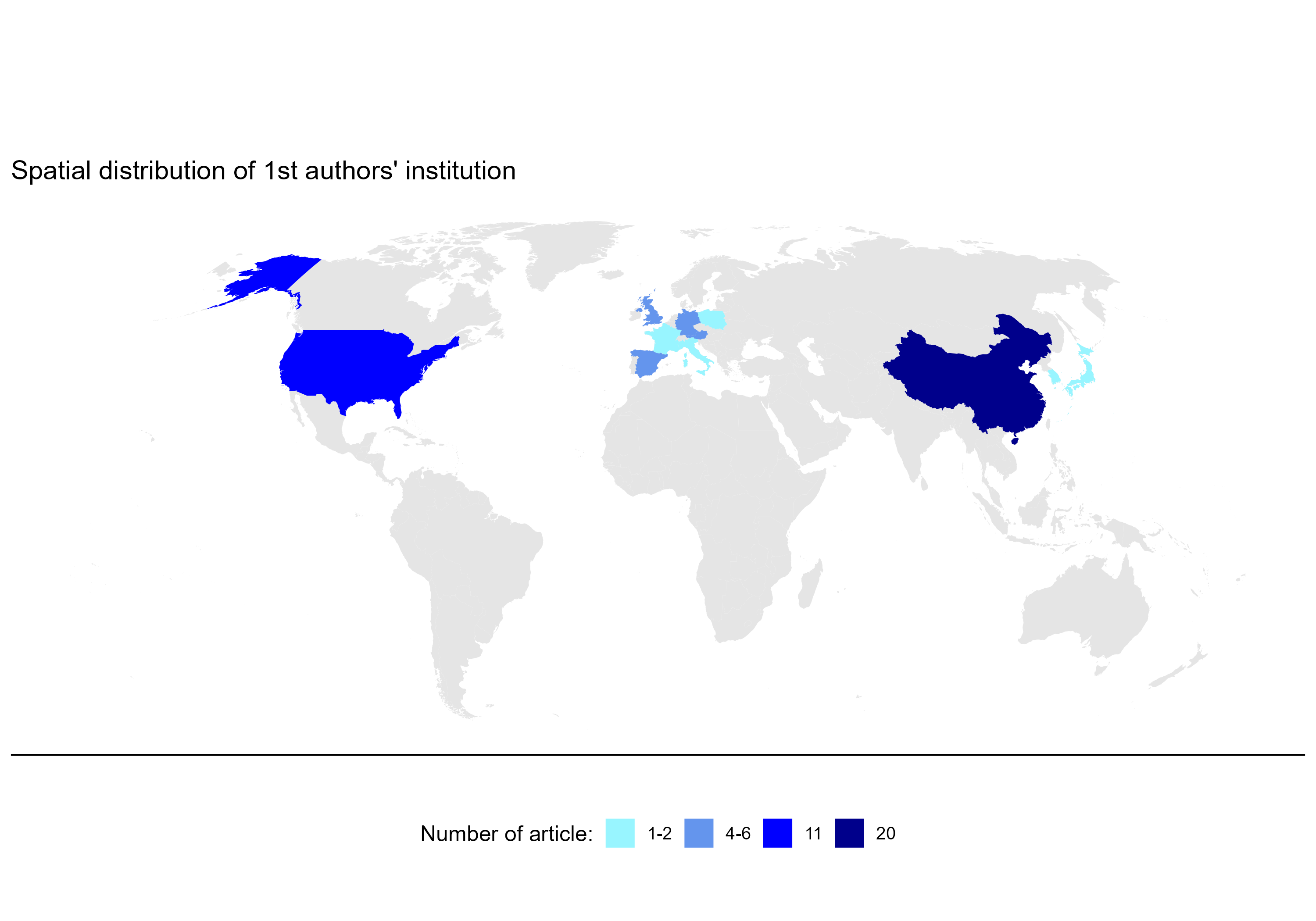
